# CA125 and age-based models for ovarian cancer detection in primary care: a population-based external validation study

**DOI:** 10.1101/2025.03.23.25324469

**Authors:** Kirsten D. Arendse, Fiona M. Walter, Gary Abel, Brian Rous, Willie Hamilton, Emma J. Crosbie, Garth Funston

**Affiliations:** Wolfson Institute of Population Health, Queen Mary University of London Barts and The London School of Medicine and Dentistry, London, United Kingdom; Primary Care Unit, Department of Public Health and Primary Care, University of Cambridge, United Kingdom; University of Exeter Medical School, University of Exeter; Cambridge University Hospitals NHS Foundation Trust, Cambridge; Gynaecological Oncology Research Group, Division of Cancer Sciences, University of Manchester; Manchester Academic Health Sciences Centre, Department of Obstetrics and Gynaecology, Manchester University NHS Foundation Trust

**Keywords:** Cancer antigen 125, early diagnosis, ovarian cancer, primary care, predictive model, validation

## Abstract

**Background:** Cancer antigen-125 (CA125) is widely used to investigate symptoms of possible ovarian cancer (OC) in primary care. However, cancer risk varies with age as well as CA125 level. We externally validated the Ovatools models, which provide CA125- and age-specific OC risk.

**Methods:** The performance of Ovatools in predicting OC diagnosis within 12 months of primary care CA125 was examined using English healthcare data for women <50 and ≥50 years. Discrimination and calibration were examined, accuracy was calculated at varying risk thresholds and compared to CA125 ≥35U/ml. We estimated OCs missed/detected by Ovatools in hypothetical diagnostic pathways, including a two-threshold pathway where moderate risk (1-2.9%) triggered primary care ultrasound, and higher risk (≥3%) triggered urgent cancer referral.

**Results:** 342,278 women were included, 0.63% had OC. The AUC was 0.95 in women ≥50 and 0.89 in women <50. When sensitivity/specificity was matched to CA125 ≥35U/ml, Ovatools showed marginally improved performance across other accuracy metrics (≥50 years). In a two-threshold pathway (≥50 years), 18.3% identified for urgent referral and 1% identified for ultrasound had OC.

**Discussion:** Ovatools performed well on external validation. Ovatools could be used to support informed decision-making and to triage women for further investigation based on cancer risk.

## Introduction

Globally in 2022, there were approximately 324,000 new cases and 200,000 deaths from ovarian cancer (OC) (1). In the United Kingdom (UK), 67% of women with OC are diagnosed with advanced disease, for which 5-year survival rates are 30% and 15% for stage III and IV, respectively (2). Large trials have not demonstrated mortality benefit from screening for OC,(3,4) and most women are diagnosed following a symptomatic presentation in primary care (5). Cancer Antigen 125 (CA125) is used in many countries as the first-line test for possible OC in symptomatic women (6). CA125 has reasonable accuracy at the standard threshold (≥35U/ml) within English primary care with a Positive Predictive Value (PPV) for invasive OC of 9% (7). However, the probability of OC varies markedly by both CA125 level and age, so older women with CA125 levels just below 35U/ml are more likely to have cancer than younger women with CA125 values well above this threshold (7). For some tests, such as prostate-specific antigen (PSA), age-specific thresholds are employed in place of a single threshold and this approach has been proposed for CA125 (8,9).

The Ovatools prediction model, developed using CA125 results and age data from over 50,000 women tested in English primary care, provides the probability of OC to guide clinical decisions on the need for further investigation (10). A potential advantage of using risk models is that thresholds for further investigation can be applied in line with national guidelines, such as the 3% risk threshold used in England for urgent cancer referral recommendations, thereby facilitating timely investigation in those at higher risk. This is of particular relevance to OC, as sequential primary care tests (CA125 followed by ultrasound) are required to trigger an urgent cancer referral in England and several other countries, potentially contributing to prolonged periods of testing in primary care even in those at evidently high risk. Evidence that the most common type of OC, high-grade serous, exhibits a median early stage (I-II) pre-diagnostic clinical phase of only 12 months (11), and that treatment delays of 1 month are associated with poorer survival in OC (12) highlights the need for accurate triage approaches and streamlined diagnostic and treatment pathways.

In this study, our primary aim was to externally validate the Ovatools models in a large representative primary care population, to assess model performance and generalisability. In addition, we sought to determine diagnostic accuracy at clinically relevant risk thresholds and explore potential implications for cancer detection when using different risk thresholds to guide further investigation or referral within primary care in England.

## Methods

### Study design and data sources

This was a retrospective cohort study using English primary care data from the Clinical Research Practice Datalink (CPRD) Aurum dataset and linked cancer registry data from the National Cancer Registration and Analysis Service (NCRAS) (13,14). CPRD Aurum comprises anonymised, coded, electronic patient health records from GP surgeries which use the EMIS clinical software (15) and is broadly representative of the UK population (16). They include data on demographics, laboratory investigations, prescriptions, ethnicity and deprivation. NCRAS collects data on all patients in England diagnosed with cancer, including incidence date, histology, morphology, and stage at diagnosis. GP practices included in the model development study were excluded from the external validation dataset.

### Participants

We applied the same criteria used in the model development study (7) when defining the cohort but included more contemporaneous data (data up to 2017 rather than up to 2014). We included women with a valid CA125 measurement recorded in CPRD between 1 May 2011 and 31 December 2017. The first CA125 test recorded during this period was considered the *index test*. Women aged <18 years on the date of their index test, those with a previous CA125 test in the year before their index test and those with a previous diagnosis of any OC (including borderline ovarian tumours) were excluded. Only CA125 values recorded in standard units (U/ml, IU/ml, KU/L, KIU/ml) were included. CA125 entries were considered invalid if the value was missing, zero or below zero.

### Clinical outcomes

The primary clinical outcome used for evaluating model performance was invasive OC recorded in NCRAS within 12 months of the index CA125 test. Invasive OC was defined using the International Classification of Diseases (ICD)-10 codes by the World Health Organization and Federation of International Gynaecology and Obstetrics and included ovarian malignancy (C56), fallopian tube malignancy (C57.0), and primary peritoneal malignancy (C48.1, C48.2). Borderline ovarian tumours/neoplasms of uncertain behaviour of the ovary (D39.1) were excluded from the primary outcome definition. Given changes in the coding of borderline ovarian tumours over time, ICD-02 and ICD-03 tumour morphology and histology codes were reviewed in consultation with a clinical pathologist (BR) to ensure appropriate classification (Supplement 1). A sub-analysis was performed with early-stage (I-II) invasive OC as the outcome. We separately evaluated a second predictive model which used any OC as the outcome including both borderline ovarian tumours and invasive OC in the outcome definition.

### Descriptive and demographic variables

Deprivation was measured at GP practice level using Townsend deprivation scores, a measure of socioeconomic deprivation (17). Townsend scores were grouped into quintiles, with quintile one being the least- and five being the most deprived. Ethnicity was categorised based on codes within CPRD into five groups in line with Office for National Statistics definitions: (1) Asian or Asian British, (2) Black or Black British, (3) Mixed, (4) Other, and (5) White or White British (18). Only the year of birth is recorded in CPRD to protect patient anonymity, therefore a birthday and month of 1 July was assigned to all patients to derive age at index test.

### Statistical analysis

#### Estimating the risk of ovarian cancer

The Ovatools prediction models were originally developed using logistic regression and incorporated continuous CA125 level and continuous age, transformed using restricted cubic splines to account for non-linear relationships between variables. Separate models exist to predict the risk of (i) invasive OC and (ii) any OC. Full model specifications have previously been published (10). For external validation in this study, we applied the prespecified models, using the same Knot placements for CA125 and age, to the validation dataset. We used logistic regression to determine individuals’ log odds of invasive OC (Supplement 2), which were converted and reported as probability [0 to 1]. This was repeated for the any OC model. The hypothetical predicted risk of invasive OC and any OC that would occur for all ages between 18 and 89 years (using age in years as a continuous variable) at all CA125 levels between 1-1000U/ml have been made available (19).

To simplify Ovatools use in practice, we also estimated mean predicted risks by CA125 level (1-1000U/ml) and age group (18-29 years, 30-39 years, 40-49 years, 50-59 years, 60-69 years, 70-79 years and 80-89 years) (20). For example, two women aged 39 and 35 years with CA125 results of the same value would have the same predicted risk because they fall within the same age group. We report the closest integer CA125 values (in U/ml) that equated to average Ovatools risks of ∼1% and ∼3% for each age group (Supplement 3) to demonstrate possible CA125 thresholds for pelvic ultrasound/urgent cancer referral by age group. These thresholds were chosen for examination in this study as ≥1% risk of cancer is often used by the National Institute for Health and Care Excellence (NICE) when recommending primary care tests in symptomatic patients, such as chest X-ray for possible lung cancer, and ≥3% as this threshold is used when recommending urgent cancer pathway referral (21).

#### External model validation

To assess Ovatools model performance using risk predictions by age (continuous) and risk predictions by age group, discrimination and calibration metrics were calculated. Discrimination is the ability to differentiate between those who experienced an event, here, invasive OC or any OC, from those who did not (22), and was determined by measuring the area under the curve (AUC). Calibration measures how closely predicted risk aligns with the proportion of those experiencing an outcome (23). Mean calibration (calibration-in-the-large, CITL) and calibration slopes were provided when constructing calibration plots using the Stata package, *pmcalplot* (24,25). We used the provided slope measurements to calculate the intercept. Models with an intercept close to 0 and a slope close to 1 were considered well-calibrated. Good calibration is most important at risk levels close to potential clinical decision thresholds (1% and 3%). Therefore, we performed an additional analysis where participants with a predicted risk level >5% (1.7% of the cohort) were excluded from the calibration plot. The mean predicted risk was plotted against the mean outcome by risk level and displayed graphically (Supplement 4.1). We also assessed for variation in the models’ performance using risk predictions for continuous age for the following demographics: (i) age (comparing women <50 and ≥50 years), (ii) ethnicity groups, and (iii) deprivation quintiles. The mean predicted risk was plotted against the mean outcome for each subgroup with demographic variable categories and performance metrics measured (AUC, slope, and intercept). We assessed the performance of the invasive OC model (using continuous age) with early-stage invasive OC as the outcome (i.e. excluding missing stage, and stage III-IV), as well as the any OC model (including borderline ovarian tumours).

#### Diagnostic accuracy

Diagnostic accuracy was calculated using the Stata package, *diagt* (26), with the PPV, negative predictive value (NPV), sensitivity and specificity reported with 95% confidence intervals (CI) (27). We calculated the diagnostic accuracy of Ovatools to predict invasive OC using both predicted risk by continuous age and age group and compared this to using CA125 ≥35U/ml. We measured the accuracy of several example thresholds for Ovatools, including at ≥1% and ≥3% predicted risk, and compared this to CA125 at ≥35U/ml. We also calculated the accuracy of Ovatools using the risk level with the same sensitivity as CA125 ≥35U/ml. The accuracy of Ovatools using risk predictions by continuous age was compared by demographic categories, (i) age (above and below 50 years), (ii) ethnicity and (iii) deprivation quintiles. We also report the diagnostic accuracy of using Ovatools predicted risk by CA125 level and age group (Supplement 4.5).

#### Clinical implications

We estimated the number of women who had a CA125 test per year in England based on CPRD data and published population statistics (28,29) (Supplement 5), and used the Ovatools accuracy metrics to approximate how many false/true positives and negatives would occur based on several exemplar pathways including one in which a 1%-2.9% risk triggers primary care ultrasound and ≥3% risk triggers urgent cancer referral. This was calculated separately for women <50 and ≥50 years, as well as when using risk predictions by age group. All data management and analyses were conducted in Stata version 18.0 (30).

### Sample size considerations

We calculated sample size requirements for precise estimation of observed divided by expected (O/E) cases, calibration slope, the C-statistic and net benefit at a referral threshold of 3%, using inputs from the development study and following guidance by Riley *et al*. (31), who recommend the sample size is at least as large as the maximum of the four required figures (Supplement 6). The largest of these values was 226,968 subjects.

### Patient and Public Involvement

Input was obtained from a patient and public involvement (PPI) group, some of whose members had experience with CA125 testing and OC. Preliminary study findings were shared and views on key findings and potential implications of the work for patients and the public were obtained and informed manuscript preparation. The PPI group will contribute to the dissemination of study findings.

## Results

### Participant characteristics

After applying the exclusion criteria, 342,278 participants were included in the study (Supplement 8). The median age was 53 years (interquartile range: 44-66). Most participants were categorised as White or White British (85%). Within 12 months of index CA125, 2,143 (0.63%) women were diagnosed with invasive OC and 2,655 (0.78%) with any OC (**Table 1**). Stage data was missing for 15% of women with invasive OC: of those with a recorded stage, 1,247 (68%) had advanced disease (Stage III or IV). Most invasive OCs were of epithelial origin (92%) (Supplement 9).

**Table 1:**
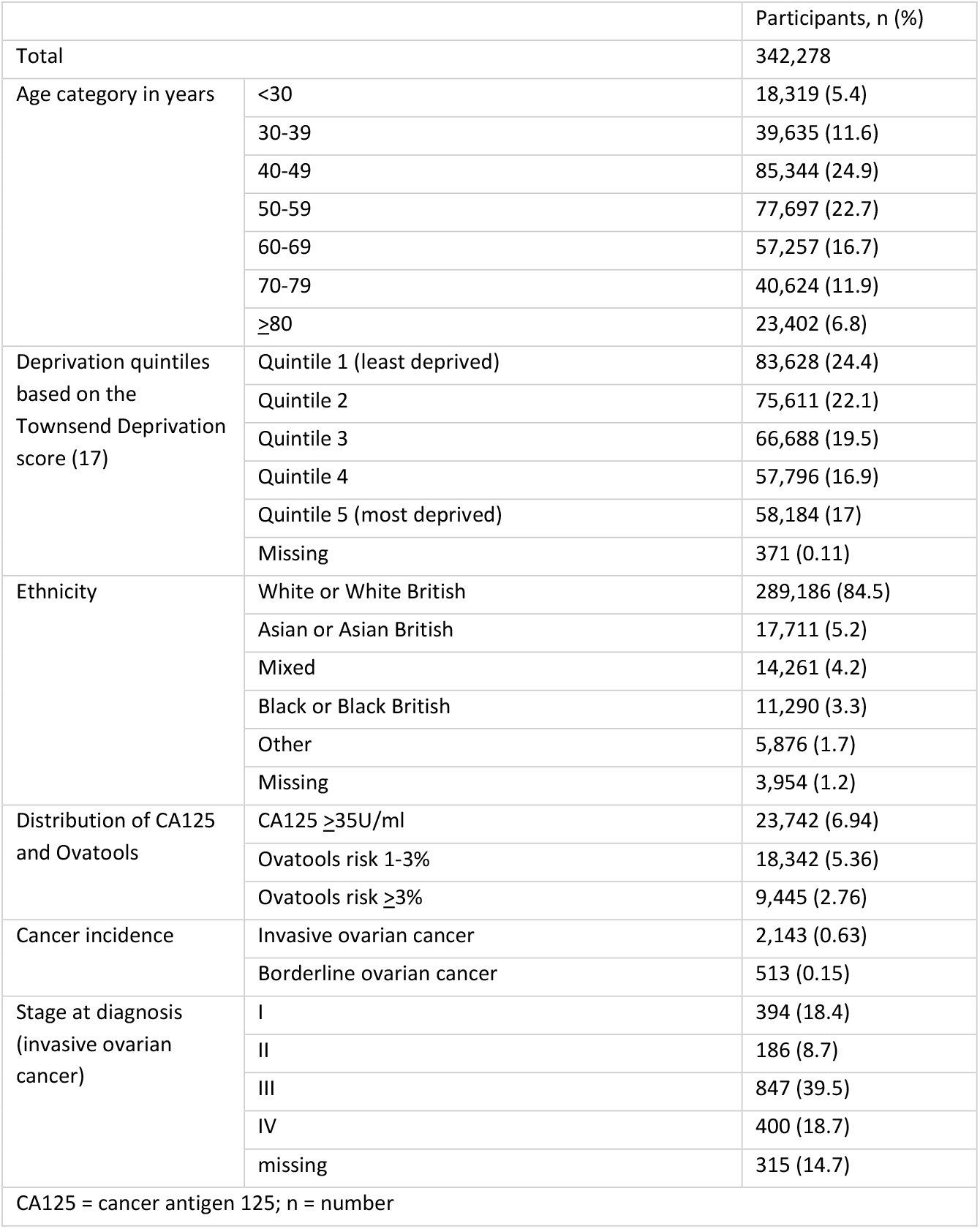
Cohort characteristics, cancer incidence, stage, and distribution of CA125 and Ovatools.

|  |  | Participants, n (%) |
| --- | --- | --- |
| Total |  | 342,278 |
| Age category in years | <30 | 18,319 (5.4) |
|  | 30-39 | 39,635 (11.6) |
|  | 40-49 | 85,344 (24.9) |
|  | 50-59 | 77,697 (22.7) |
|  | 60-69 | 57,257 (16.7) |
|  | 70-79 | 40,624 (11.9) |
|  | ≥80 | 23,402 (6.8) |
| Deprivation quintiles based on the Townsend Deprivation score (17) | Quintile 1 (least deprived) | 83,628 (24.4) |
|  | Quintile 2 | 75,611 (22.1) |
|  | Quintile 3 | 66,688 (19.5) |
|  | Quintile 4 | 57,796 (16.9) |
|  | Quintile 5 (most deprived) | 58,184 (17) |
|  | Missing | 371 (0.11) |
| Ethnicity | White or White British | 289,186 (84.5) |
|  | Asian or Asian British | 17,711 (5.2) |
|  | Mixed | 14,261 (4.2) |
|  | Black or Black British | 11,290 (3.3) |
|  | Other | 5,876 (1.7) |
|  | Missing | 3,954 (1.2) |
| Distribution of CA125 and Ovatoools | CA125 ≥35U/ml | 23,742 (6.94) |
|  | Ovatoools risk 1-3% | 18,342 (5.36) |
|  | Ovatoools risk ≥3% | 9,445 (2.76) |
| Cancer incidence | Invasive ovarian cancer | 2,143 (0.63) |
|  | Borderline ovarian cancer | 513 (0.15) |
| Stage at diagnosis (invasive ovarian cancer) | I | 394 (18.4) |
|  | II | 186 (8.7) |
|  | III | 847 (39.5) |
|  | IV | 400 (18.7) |
|  | missing | 315 (14.7) |
| CA125 = cancer antigen 125; n = number |  |  |

### Model External Validation

Using predicted risk by continuous age, the invasive OC model showed good overall discrimination, with an AUC of 0.946 (95% CI: 0.939-0.953) (**Table 2**). The mean calibration was close to 0, the calibration slope was close to 1 and the intercept was close to 0, suggesting minimal overfitting. Model performance was tested by predicted risk level, and mean calibration was acceptable at thresholds of interest (∼1% and ∼3% risk) but there was less agreement between observed outcomes and predicted outcomes at extreme risk levels (Supplement 4.1). Performance was better for women ≥50 years (AUC 0.951, 95% CI: 0.942-0.960) than those <50 years (AUC 0.889, 95% CI: 0.886-0.892). The model performed well across deprivation levels and ethnicity groups (Supplement 4.2). When applying early-stage OC as the outcome, model performance was slightly lower (AUC 0.885, Intercept 0.0038, Slope 0.751). When using the any OC model including border tumours in the outcome, there was good calibration but marginally poorer discrimination (AUC 0.925, 95% CI: 0.918-0.931) than for the invasive OC model (Supplement 4.3). When using risk predictions by age group, the invasive OC model performed well, with only marginally lower metrics than when using risk predictions by continuous age (Supplement 4.4). Performance using age-group-based CA125 thresholds showed a similar pattern across ages compared to using the model with continuous age, with a better AUC for women ≥50 years (0.933, 95% CI: 0.922-0.943), compared to those <50 years (AUC 0.872, 95% CI: 0.842-0.901).

**Table 2:**
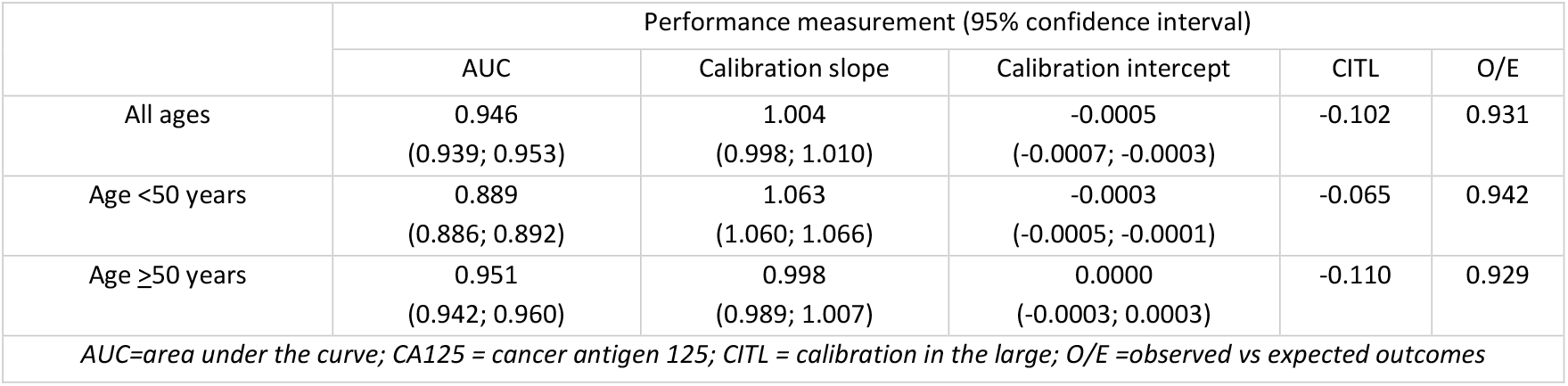
Performance of the Ovatools invasive ovarian cancer model (using continuous age) on external validation.

### The diagnostic accuracy of Ovatools using continuous age vs CA125 ≥35U/ml

The diagnostic accuracy of Ovatools for invasive OC using continuous age is shown at risk thresholds ≥1% and ≥3% and compared to CA125 at ≥35U/mL for women <50 and ≥50 years (**Table 3**). For women ≥50 years, Ovatools at ≥1% had a higher sensitivity than CA125 at >35U/ml (91.1% vs 86.5%) but lower PPV (7.2% vs 12.5%) and specificity (89.1% vs 94.3%). For women <50 years, Ovatools at ≥1% had a lower sensitivity than CA125 >35U/ml (61.9% vs 75.3%), with a greater specificity (96.9% vs 92.5%) and PPV (4.0% vs 2.0%). All diagnostic accuracy metrics for both Ovatools and CA125 ≥35U/ml were greater among women ≥50 than <50 years. When using the Ovatools risk thresholds that had the same sensitivity and specificity as CA125 ≥35U/ml, Ovatools showed modest improvement across other accuracy metrics for women ≥50 years but not for women <50 years (**Table 3**). For women ≥50 years, an Ovatools threshold of ≥2.12% had the same sensitivity (86.5%) as CA125 at ≥35U/ml, but a greater specificity (94.8% vs 94.3%) and PPV (13.4% vs 12.5%). For women with early-stage invasive OC, Ovatools at ≥1% had a higher sensitivity than CA125 >35U/ml (70.7% vs 66.7%) but lower specificity (92.4% vs 93.6%) and a similar PPV (1.6% vs 1.7%) (Supplement 10).

**Table 3:** The diagnostic accuracy of Ovatools using predictions by continuous age at multiple risk thresholds, and CA125 at ≥35U/ml.

| Sub-group<br>(invasive ovarian cancer incidence) | CA125 or Ovatoools threshold | Sensitivity % (95% CI) | Specificity % (95% CI) | PPV % (95% CI) | NPV % (95% CI) |
| --- | --- | --- | --- | --- | --- |
| Age $< 50$ years<br>(0.21%) | CA125 $\geq 35$U/ml | 75.3 (70.0; 80.0) | 92.5 (92.3; 92.6) | 2.0 (1.8; 2.3) | 99.9 (99.9; 100) |
| | $\geq 1\%$ | 61.9 (56.1; 67.4) | 96.9 (96.8; 97.0) | 4.0 (3.5; 4.7) | 99.9 (99.9; 99.9) |
| | $\geq 3\%$ | 45.2 (39.4; 51.0) | 99.4 (99.3; 99.4) | 12.7 (10.8; 14.9) | 99.9 (99.9; 99.9) |
| | $\geq 0.52\%^a$ | 75.3 (70.0; 80.0) | 91.6 (91.5; 91.7) | 1.8 (1.6; 2.1) | 99.9 (99.9; 100) |
| | $\geq 0.57\%^b$ | 73.6 (68.2; 78.5) | 92.5 (92.4; 92.7) | 2.0 (1.8; 2.3) | 99.9 (99.9; 100) |
| Age $\geq 50$ years<br>(0.93%) | CA125 $\geq 35$U/ml | 86.5 (84.8; 88.0) | 94.3 (94.2; 94.4) | 12.5 (11.9; 13.1) | 99.9 (99.8; 99.9) |
| | $\geq 1\%$ | 91.1 (89.7; 92.4) | 89.1 (88.9; 89.2) | 7.2 (6.9; 7.6) | 99.9 (99.9; 99.9) |
| | $\geq 3\%$ | 83.1 (81.3; 84.8) | 96.5 (96.4; 96.6) | 18.3 (17.5; 19.1) | 99.8 (99.8; 99.9) |
| | $\geq 2.12\%^c$ | 86.5 (84.8; 88.0) | 94.8 (94.7; 94.9) | 13.4 (12.8; 14.1) | 99.9 (99.8; 99.9) |
| | $\geq 1.95\%^d$ | 87.7 (86.1; 89.1) | 94.3 (94.2; 94.4) | 12.5 (11.9; 13.1) | 99.9 (99.9; 99.9) |
| CA125 = cancer antigen 125, CI = confidence interval; NPV = negative predictive value; PPV = positive predictive value<br>The following Ovatoools thresholds were selected because they had the same sensitivity<sup>a,c</sup> or specificity<sup>b,d</sup> as CA125 $\geq 35$U/ml. | | | | | |

### CA125 levels equating to Ovatools risk ∼1% and ∼3% by age group

The CA125 levels equating to Ovatools risk levels of ∼1% and ∼3% for invasive OC by age group are displayed in **Figure 1** (numerical values are in Supplement 3). CA125 values were highest in women aged 30-39 years (1%: CA125=59U/ml, 3%: CA125=160U/ml) and 40-49 years (1%: CA125=58U/ml, 3%: CA125=157U/ml), and lowest in women in the 60-69 years age group (1%: CA125=22U/ml, 3%: CA125=37U/ml). The CA125 level equating to ∼1% Ovatools risk was higher than the current CA125 threshold (≥35U/ml) in age groups 30-39 years and 40-49 years but lower in all other age groups. In the 60-69 years group, the CA125 value equating to ∼3% risk was only 2U/ml higher than the current CA125 threshold for primary care ultrasound. Ovatools thresholds equating to ∼1% risk had a greater sensitivity for all age groups except for women 30-39 years and 40-49 years, when compared to CA125 at ≥35U/ml (Supplement 4.5).

**Figure 1:**
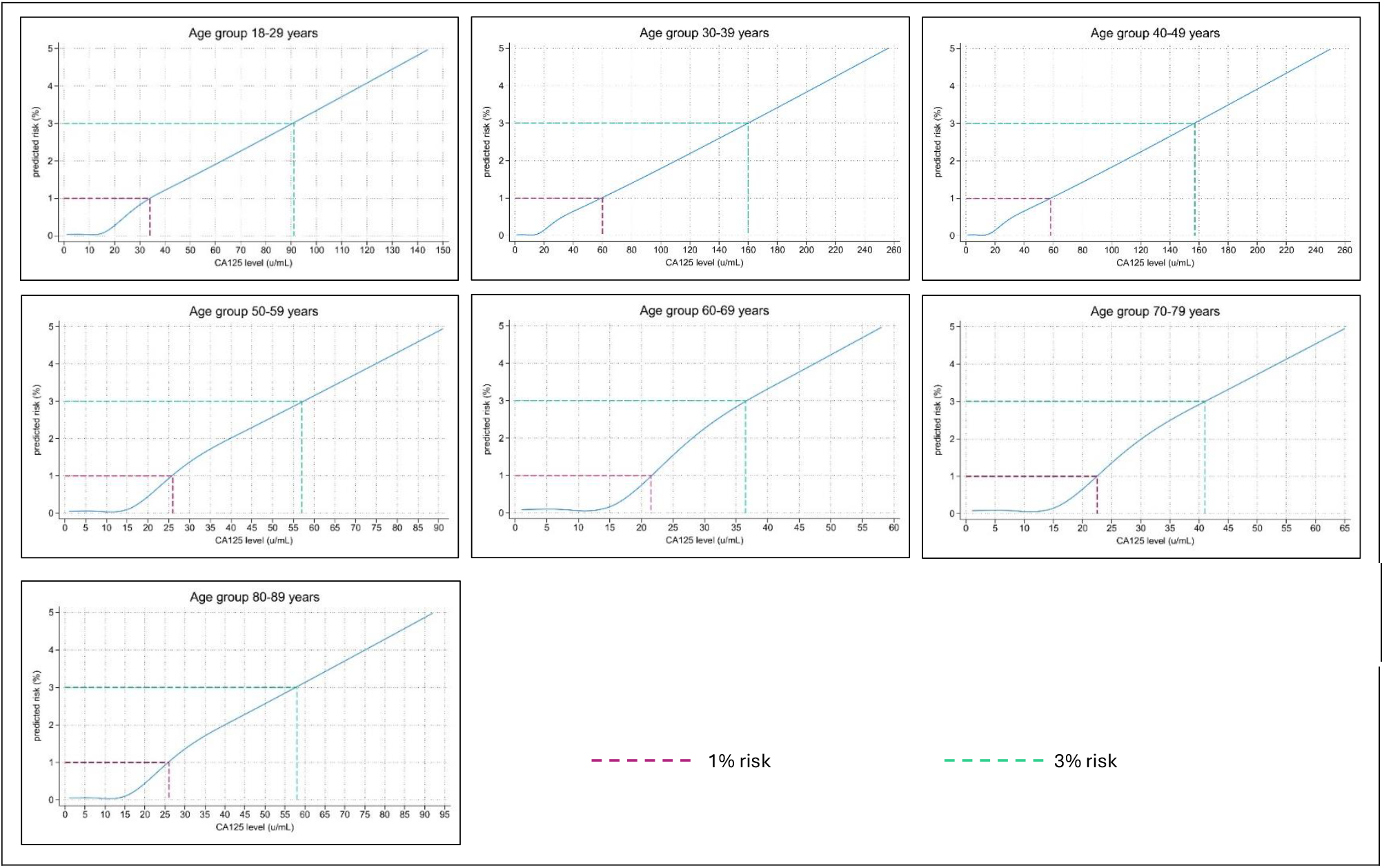
The predicted risk of invasive ovarian cancer by CA125 level and age group using Ovatools,and CA125 levels equating to ∼1% and ∼3% risk.

### Clinical implications of using Ovatools in general practice

In England in 2022, we estimated 218,335 women were tested using CA125 in primary care, with 57.7% (n=126,918) of the population being ≥50 years and 42.3% (n=91,417) being <50 years. Using standard practice, 8,137 (6.4%) of CA125-tested women ≥50 years and 7,008 (7.7%) <50 years would have a CA125 result ≥35U/ml and therefore qualify for primary care ultrasound under NICE guidance (**Figure 2** and **Table 5**). By comparison, if an Ovatools risk of 1-2.9% triggered primary care ultrasound and ≥3% triggered urgent cancer pathway referral, 14,803 (11.7%) and 2,921 (3.2%) of women ≥50 years and <50 years, respectively, would qualify for further investigation following CA125 (ultrasound or urgent cancer referral). This equates to 6,666 more women ≥50 but 4,087 fewer women <50 years being investigated further following CA125 testing (**Table 5**). This approach would result in 54 additional women with OC ≥50 years but 26 fewer women with OC <50 years being identified for investigation in England per year. If the Ovatools pathway at the proposed thresholds were used for women ≥50 years only, 1 in 5 (18.3%) of high-risk women (≥3% Ovatools risk) identified for urgent cancer referral, and 1 in 100 moderate-risk women (1-2.9% Ovatools risk) identified for primary care ultrasound would have invasive OC. For every 123 additional women ≥50 years sent for further investigation using Ovatools compared to standard practice, 1 additional case of invasive OC could be identified.

**Table 5:** Number of women sent for further testing and detected with invasive OC using CA125 ≥35U/ml and Ovatools at multiple thresholds (using continuous age).

| Age group | Threshold for primary care ultrasound | USS following CA125 | Change in pelvic ultrasounds using Ovatoools | Urgent cancer referrals (risk $\geq 3\%$) | Additional women testing further using Ovatoools | Invasive OC detected | Additional OC detected using Ovatoools | Additional OC diagnoses / 100,000 CA125-tested women |
| --- | --- | --- | --- | --- | --- | --- | --- | --- |
| $\geq 50$ years | CA125 $\geq 35$U/mL | 8,137 | NA | NA | NA | 1,017 | NA | NA |
|  | Ovatoools 1-2.9% | 9,456 | 1,319 | 5,347 | 6,666 | 1,071 | 54 | 59 |
|  | *Ovatoools 1.95-2.9% | 2,906 | -5,231 | 5,347 | 116 | 1,031 | 14 | 15 |
|  | ^Ovatoools 2.12-2.9% | 2,222 | -5,915 | 5,347 | -568 | 1,017 | 0 | 0 |
| $< 50$ years | CA125 $\geq 35$U/mL | 7,008 | NA | NA | NA | 144 | NA | NA |
|  | Ovatoools 1-2.9% | 2,244 | -4,764 | 678 | -4,086 | 118 | -26 | -28 |

CA125 = cancer antigen 125; OC = ovarian cancer; Ovatoools threshold with a matched specificity\* and sensitivity^ for women $\geq 50$ years.

**Figure 2:**
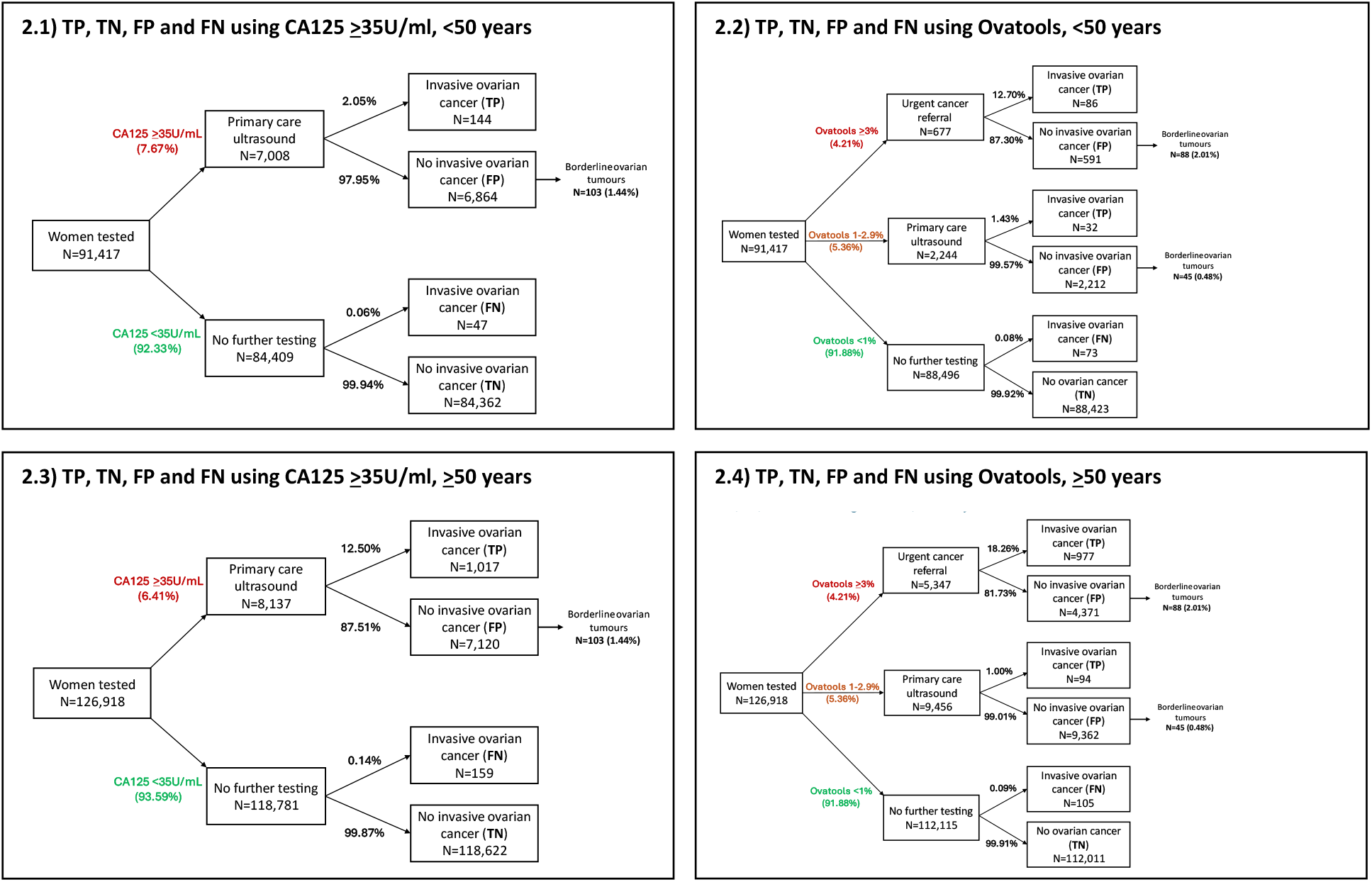
Estimated true positive (TP), false positive (FP), false negative (FN) and true negative (TN) cases that could occur in England per year using the CA125 ≥35U/ml to trigger primary care ultrasound compared to using Ovatools at 1-2.9% risk to trigger primary care ultrasound and ≥3% to trigger urgent cancer referral for women <50 years (2.1 & 2.2), and women ≥50 years (2.3 & 2.4).

**Table 5** demonstrates the clinical utility of using Ovatools risk thresholds with matched sensitivity (≥2.12%) and specificity (≥1.95%) to that of CA125 ≥35U/ml for women ≥50 years. For women ≥50 years, if 2.12-2.9% risk triggered primary care ultrasound, while ≥3% triggered urgent cancer referral, 568 fewer women would be sent for further investigation compared to standard practice while identifying the same number of invasive OC cases. If 1.95-2.9% risk triggered primary care ultrasound and ≥3% triggered urgent cancer referral, only 116 additional women would be investigated further while identifying 14 additional cases of invasive OC (1 in 8 women tested further). Clinical utility was repeated using age-based CA125 thresholds approximating ∼1% and ∼3% risk to trigger ultrasound for referral, respectively, showing similar findings, with an increase in OC cases detected in women ≥50 years, but additional missed cases in women <50 years (Supplement 5).

## Discussion

This study found that Ovatools, an age- and CA125-based model to predict the risk of invasive OC, performed well on external validation in 342,278 CA125-tested women in English primary care. Performance was similar across ethnic and sociodemographic groups but greater in women ≥50 than <50 years. Depending on the threshold(s) chosen, Ovatools age-adjusted risk thresholds reduced false positives or false negatives when compared to CA125 ≥35U/ml. When the sensitivity or specificity of Ovatools thresholds were matched to CA125 ≥35U/ml, Ovatools exhibited a modest improvement across other accuracy metrics (PPV, NPV) for all women ≥50 years and a reduction in primary care ultrasounds among women without OC. In a scenario where moderate Ovatools risk (1-2.9%) triggers primary care ultrasound and higher Ovatools risk (≥3%) triggers urgent cancer referral for women ≥50 years, more women would be selected for investigation in comparison to the current practice, with 1 in 123 additional women selected for further testing having invasive OC, and 1 in 5 (18.3%) of high-risk women selected for direct urgent cancer referral having invasive OC. In women <50 years, the same risk thresholds would result in fewer unnecessary referrals (false positives) but additional missed OC cases (false negatives) when compared to current practice, so alternative thresholds or diagnostic strategies may be more appropriate for this age group and warrants further investigation.

The Ovatools model enables thresholds for further investigation to be chosen based on age- and CA125-derived OC risk. Model parameters and risk levels equating to a wide range of CA125 levels and ages are published alongside this paper to enable thresholds to be set in line with local or national priorities. Results could also be used to inform individual doctor-patient choices on further investigation. Using an Ovatools threshold (≥2.12% risk) with an equivalent sensitivity to CA125 ≥35U/ml for women ≥50 years could reduce unnecessary ultrasound investigations without increasing missed OC cases. However, to achieve earlier detection, more sensitive methods to identify symptomatic OC in primary care and fast-track high-risk women through the diagnostic pathway are needed, particularly in the absence of OC screening (3). Ovatools could be used to select high-risk women for expedited investigation, such as ≥3% risk to trigger urgent cancer referral in CA125-tested women in line with the 3% NICE threshold for urgent referral of symptomatic patients for other cancers (21). This approach would enable expedited specialist gynaecological assessment and ultrasound using the gold standard Risk of Malignancy Index or IOTA for women with a high risk of undiagnosed invasive OC (32). Under current NICE guidelines, these high-risk women require a GP-requested ultrasound before urgent referral, which can take weeks or months (33), are associated with longer primary care intervals in OC (34) and are generally not interpreted using gold standard approaches. The proportion of women in the ≥3% risk Ovatools group who had invasive OC (12.7% and 18.3% of women <50 and ≥50 years, respectively) would far exceed the current gynaecological urgent cancer referral pathway conversion rate for England (2.9%) (35). In addition, offering women at low-but-not-no-risk (1-2.9%) of invasive OC ultrasound in primary care (the current standard in women with raised CA125) could reduce false negative results, which are associated with longer test-diagnostic intervals (36). Such approaches have the potential to increase timely OC detection but would rely on improved timely access to transvaginal ultrasound within primary care, which is currently planned in England as part of the expansion of community diagnostic centres (37).

Applying age-specific thresholds would have significant implications for patients and the healthcare service. Therefore, the feasibility, acceptability and cost-effectiveness of any change should be understood prior to clinical implementation. Further, while CA125 is used as the first-line test in England and several other countries, elsewhere CA125 and ultrasound (or computed tomography) are used in parallel and different Ovatools risk thresholds may be more appropriate for those settings (6). While Ovatools risk at individual ages and CA125 levels could be readily incorporated into blood test reports to inform individual decision-making, we evaluated the CA125 levels equating to clinically relevant thresholds by age group as these may be easier to implement. Similar approaches have been used for other tests such as PSA, with NICE guidelines recently updated to recommend referral at different PSA levels in different age groups (8).

Few studies have examined the performance of CA125 or OC prediction models in primary care in the UK. In the current study, CA125 (≥35U/ml) had the same sensitivity and specificity to detect invasive OC as at model development; however, we demonstrated a slightly lower PPV (7.7% vs 8.8%), and lower incidence of any OC (0.78% vs 0.93%). As in the development study (10), both CA125 and Ovatools performed less well in women <50 years, likely due to the lower OC prevalence in this group, differences in OC histology (38) and greater incidence of benign conditions which can elevate CA125. We found that a high proportion (42%) of CA125 tests were performed in women <50, but the incidence of OC was five times lower than in the group ≥50 years, while only 2% of women <50 years with a CA125 ≥35U/ml had an OC. Given the limited accuracy of CA125 and Ovatools in younger women, other tests could be considered, such as HE4, a biomarker shown to have high sensitivity to detect OC when used alongside CA125 in symptomatic women <50 years (39).

### Strengths and limitations

This study used a large dataset which was broadly representative of the English population, with ethnicity and deprivation distributions similar to national estimates (40,41). Laboratory results are automatically recorded in CPRD, leading to high levels of completeness. We were unable to determine the reasons for CA125-testing due to the use of routine data. However, our results are based on CA125 tests done on women within English primary care and therefore provide an indication of model performance in real-world clinical practice. We examined variation in performance by key demographic factors but did not include some variables (such as symptoms) previously reported to affect CA125 level and OC risk, as these did not improve performance at model development (10). Further, complex prediction models are more challenging to implement, and few are routinely used in primary care. The completeness and accuracy of NCRAS data used to determine outcomes are high (42). However, stage data was missing for 15% of invasive OCs, limiting analysis by stage. Not all women had a reference standard test and instead we rely on OC diagnoses within 12 months of CA125. Some women may be diagnosed beyond this interval, and some may develop cancer during the period following CA125 testing which may introduce bias (43). A 12-month period has been widely used in similar research (7,36) and was chosen as a compromise between minimising the inclusion of new cancers and maximizing the inclusion of relevant cancers.

## Conclusion

Ovatools performs well in identifying invasive OC in CA125-tested women, particularly in women ≥50 years. The model could be used to interpret CA125 levels within primary care and select higher-risk women for further investigation and referral. This approach has the potential to expedite diagnosis, but further work is needed to understand the feasibility, acceptability and the cost and benefits of using Ovatools within diagnostic pathways.

## Supporting information

Ovarian tumour morphology codes

Ovatools risk prediction formulae

CA125 levels equating 1% and 3% risk by age group

Additional performance metrics - by risk level, demographics and any OC

Calculating the clinical utility of Ovatools

Sample size calculation

TRIPOD checklist

Flow diagram demonstrating the application of inclusion and exclusion criteria

Ovarian tumour behaviour and morphology

Additional diagnostic accuracy metrics - by age group, cancer stage and for any OC

## Abbreviations

AUC: Area under the curve
CA125: Cancer antigen 125
CI: Confidence interval
CITL: Calibration-in-the-large
CPRD: Clinical Research Practice Datalink
ICD: International Classification of Diseases
GP: General practitioner
NCRAS: National Cancer Registration and Analysis
NICE: National Institute for Health and Care Excellence
NPV: Negative predictive value
OC: Ovarian cancer
O/E: Observed divided by expected cases
PPV: Positive predictive value
PSA: Prostate specific antigen
ROC: Receiver operating curve
SE: Standard error
TRIPOD: Transparent Reporting of a Multivariable Prediction Model for Individual Prognosis or Diagnosis
UK: United Kingdom
U/ml: Units per millilitre

## Additional Information

## Acknowledgements

We acknowledge and thank the NHS and patients, whose data was collected as part of routine care and support.

## Authors’ contributions

Conceptualisation: KDA, GF & FWM; Data curation: KDA; Analysis: KDA, GF, & GA; Funding acquisition: GF & FWM; Investigation: KDA; Methodology: KDA & GF; Supervision: GF; Writing – original draft: KDA; Writing – review and editing: KDA, GF, FWM, GA, BR, WH, & EJC.

## Ethics

The study was approved by the Independent Scientific Advisory Committee (ISAC) for the Medicines and Healthcare Products Regulatory Agency (protocol number 21_001655). All data were provided to researchers in an anonymised form, and individual consent was not required.

## Data availability

The data used for this study were provided by CPRD and NCRAS and are subject to a licensing agreement that prohibits sharing outside the research team. Data can be requested through CPRD. All Stata scripts and code lists used to clean and analyse the data will be made available on the Queen Mary Research Online, an online data repository, or as supplementary materials.

## Reporting

This study is reported in line with the Transparent Reporting of a Multivariable Prediction Model for Individual Prognosis or Diagnosis recommendations (Supplement 7) (44).

