## Supplementary material for "CA125 and age-based models for ovarian cancer detection in primary care: a population-based external validation study": Ovarian tumour morphology codes

### Supplement 1: ICD-02 / 03 morphological classification of ovarian tumours

| morphology code | behaviour | category | description |
|---|---|---|---|
| 80001 | borderline | unknown | neoplasm, uncertain whether benign or malignant |
| 80011 | borderline | unknown | tumour cells uncertain whether benign or malignant |
| 80101 | borderline | epithelial (other) | epithelial tumour, uncertain behaviour |
| 80105 | borderline | epithelial (unknown) | carcinoma in situ with microinvasion |
| 81405 | borderline | epithelial (unknown) | adenoma |
| 82401 | borderline | non-epithelial | carcinoid tumour nos |
| 83131 | borderline | epithelial (clear cell) | clear cell adenofibroma of borderline malignancy |
| 83801 | borderline | epithelial (endometrioid) | endometrioid adenoma, borderline malignancy |
| 83811 | borderline | epithelial (endometrioid) | endometrioid adenofibroma borderline malignancy |
| 84401 | borderline | epithelial (unknown) | cystadenocarcinoma borderline malignancy |
| 84411 | borderline | epithelial (serous) | serous cystadenoma, borderline malignancy |
| 84415 | borderline | epithelial (serous) | serous cystadenoma nos |
| 84421 | borderline | epithelial (serous) | serous borderline tumor, nos |
| 84422 | borderline | epithelial (serous) | borderline serous tumour with high grade dysplasia |
| 84423 | borderline | epithelial (serous) | serous cystadenoma, borderline malignancy |
| 84425 | borderline | epithelial (serous) | serous cystadenoma, microinvasion |
| 84441 | borderline | epithelial (clear cell) | clear cell cystic tumor of borderline malignancy |
| 84511 | borderline | epithelial (unknown) | papillary cystadenoma, borderline malignancy |
| 84513 | borderline | epithelial (unknown) | papillary cystadenoma borderline malignancy |
| 84515 | borderline | epithelial (unknown) | papillary cystadenoma, borderline malignancy |
| 84601 | borderline | epithelial (serous) | papillary serous cystadenoma borderline malignancy |
| 84605 | borderline | epithelial (serous) | papillary serous cystadenoma, nos |
| 84611 | borderline | epithelial (serous) | serous surface papilloma borderline malignancy |
| 84621 | borderline | epithelial (serous) | serous papillary cystic tumor of borderline malignancy |
| 84623 | borderline | epithelial (serous) | papillary serous cystadenoma, borderline malignancy |
| 84625 | borderline | epithelial (serous) | papillary serous cystadenoa, microinvasion |
| 84631 | borderline | epithelial (serous) | serous surface papillary tumor of borderline malignancy |
| 84701 | borderline | epithelial (mucinous) | cystadenoma mucinous b/l |
| 84705 | borderline | epithelial (mucinous) | mucinous cystadenoma |
| 84721 | borderline | epithelial (mucinous) | mucinous cystic tumor of borderline malignancy |
| 84722 | borderline | epithelial (mucinous) | mucinous cystic tumor of borderline malignancy with intraepithelial carcinoma |
| 84723 | borderline | epithelial (mucinous) | mucinous cystadenoma, borderline malignancy |
| 84725 | borderline | epithelial (mucinous) | mucinous cystadenoma, microinvasive |
| 84731 | borderline | epithelial (mucinous) | papillary mucinous cystadenoma borderline malignan |
| 84733 | borderline | epithelial (mucinous) | papillary mucinous cystadenoma, borderline malignancy |
| 84741 | borderline | epithelial (other) | seromucinous borderline tumour |
| 84801 | borderline | epithelial (mucinous) | low grade appendiceal mucinous neoplasm |
| 84811 | borderline | epithelial (mucinous) | mucin-producing adenocarcinoma borderline malignan |
| 85901 | borderline | non-epithelial (sex cord stromal) | sex cord stromal tumour |
| 86211 | borderline | non-epithelial (sex cord stromal) | granulosa cell theca-cell tum |
| 86221 | borderline | non-epithelial (sex cord stromal) | granulosa cell tumor, juvenile (not testis) |
| 86311 | borderline | non-epithelial (sex cord stromal) | sertoli-leydig cell tumor of intermediate differentiation |
| 86321 | borderline | non-epithelial (sex cord stromal) | gynandroblastoma |
| 86401 | borderline | non-epithelial (sex cord stromal) | sertoli cell tumour |
| 86501 | borderline | non-epithelial (sex cord stromal) | leydig cell tumour nos |
| 88101 | borderline | non-epithelial (sex cord stromal) | cellular fibroma |
| 88971 | borderline | non-epithelial | smooth muscle tumour of uncertain malignant potent |

| | | | |
|---|---|---|---|
| 89351 | borderline | non-epithelial (sex cord stromal) | stromal tumor, nos |
| 89901 | borderline | non-epithelial | mesenchymoma, nos |
| 90001 | borderline | epithelial (other) | brenner tumor, borderline malignancy |
| 90003 | borderline | epithelial (other) | brenner tumor, malignant |
| 90131 | borderline | epithelial (unknown) | adenofibroma borderline malignancy |
| 90141 | borderline | epithelial (serous) | serous adenofibroma of borderline malignancy |
| 90151 | borderline | epithelial (mucinous) | mucinous adenofibroma of borderline malignancy |
| 90731 | borderline | non-epithelial (germ cell) | gonadoblastoma |
| 90801 | borderline | non-epithelial (germ cell) | teratoma |
| 90911 | borderline | non-epithelial (germ cell) | strumal carcinoid |
| 91101 | borderline | epithelial (other) | mesonephric tumour, nos |
| 80003 | invasive | unknown | neoplasm, malignant |
| 80009 | invasive | epithelial (other) | neoplasm, malignant unknown if primary or metastatic |
| 80043 | invasive | epithelial (unknown) | malignant tumor, spindle cell type |
| 80053 | invasive | epithelial (clear cell) | malignant tumor, clear cell type |
| 80103 | invasive | epithelial (unknown) | carcinoma nos |
| 80133 | invasive | epithelial (other) | large cell neuroendocrine carcinoma |
| 80203 | invasive | epithelial (other) | carcinoma, undifferentiated nos |
| 80213 | invasive | epithelial (unknown) | carcinoma anaplastic type |
| 80223 | invasive | epithelial (unknown) | pleomorphic carcinoma |
| 80333 | invasive | epithelial (other) | pseudosarcomatous carcinoma |
| 80413 | invasive | epithelial (other) | small cell carcinoma nos |
| 80443 | invasive | epithelial (other) | small cell carcinoma, hypercalcaemic type |
| 80463 | invasive | epithelial (unknown) | non-small cell carcinoma |
| 80503 | invasive | epithelial (unknown) | papillary carcinoma nos |
| 80523 | invasive | epithelial (other) | papillary squamous cell carcinoma |
| 80703 | invasive | epithelial (other) | squamous cell carcinoma nos |
| 80713 | invasive | epithelial (other) | squamous cell carcinoma, keratinizing, nos |
| 80733 | invasive | epithelial (other) | squamous cell carcinoma small cell non-keratinisin |
| 81203 | invasive | epithelial (other) | transitional cell ca nos |
| 81403 | invasive | epithelial (unknown) | adenocarcinoma nos |
| 81409 | invasive | epithelial (unknown) | adenocarcinoma, nos unknown if primary or metastatic |
| 81443 | invasive | epithelial (unknown) | adenocarcinoma intestinal type |
| 82403 | invasive | non-epithelial | carcinoid tumor nos |
| 82433 | invasive | non-epithelial | goblet cell carcinoid |
| 82463 | invasive | epithelial (other) | neuroendocrine carcinoma |
| 82493 | invasive | non-epithelial | atypical carcinoid/grade 2 neuroendocrine tumour |
| 82553 | invasive | epithelial (unknown) | adenocarcinoma with mixed subtypes |
| 82603 | invasive | epithelial (unknown) | adenocarcinoma papillary nos |
| 83103 | invasive | epithelial (clear cell) | clear cell adenocarcinoma nos |
| 83203 | invasive | non-epithelial (sex cord stromal) | granular cell carcinoma |
| 83233 | invasive | epithelial (other) | mixed cell adenocarcinoma |
| 83803 | invasive | epithelial (endometrioid) | endometrioid carcinoma |
| 83823 | invasive | epithelial (endometrioid) | endometrioid adenocarcinoma, secretory variant |
| 84403 | invasive | epithelial (unknown) | cystadenocarcinoma nos |
| 84413 | invasive | epithelial (serous) | serous cystadenocarcinoma nos |
| 84503 | invasive | epithelial (unknown) | cystadenocarcin papillary nos |
| 84603 | invasive | epithelial (serous) | papillary serous cystadenocarcinoma |
| 84613 | invasive | epithelial (serous) | serous surface papillary carcinoma |
| 84703 | invasive | epithelial (mucinous) | mucinous cystadenocarcinoma nos |
| 84713 | invasive | epithelial (mucinous) | papillary mucinous cystadenocarcinoma |
| 84743 | invasive | epithelial (other) | seromucinous carcinoma |
| 84803 | invasive | epithelial (mucinous) | mucinous adenocarcinoma |
| 84813 | invasive | epithelial (mucinous) | mucin-producing adenocarcinoma |
| 84823 | invasive | epithelial (mucinous) | mucinous adenocarcinoma, endocervical type |

| | | | |
|---|---|---|---|
| 84903 | invasive | epithelial (mucinous) | signet ring cell carcinoma |
| 85603 | invasive | epithelial (other) | adenosquamous carcinoma |
| 85743 | invasive | epithelial (unknown) | adenocarcinoma with neuroendocrine differentiation |
| 85753 | invasive | epithelial (unknown) | metaplastic carcinoma, nos |
| 86003 | invasive | non-epithelial (sex cord stromal) | theca cell carcinoma |
| 86201 | invasive | non-epithelial (sex cord stromal) | granulosa cell tumor nos |
| 86203 | invasive | non-epithelial (sex cord stromal) | granulosa cell tumor, malignant |
| 86313 | invasive | non-epithelial (sex cord stromal) | sertoli-leydig cell tumour, poorly differentiated |
| 86503 | invasive | non-epithelial (sex cord stromal) | leydig cell tumor, malignant |
| 86703 | invasive | non-epithelial | steroid cell tumor, malignant |
| 88003 | invasive | non-epithelial | sarcoma nos |
| 88103 | invasive | non-epithelial | fibrosarcoma |
| 88513 | invasive | non-epithelial | liposarcoma, well differentiated |
| 88903 | invasive | non-epithelial | leiomyosarcoma nos |
| 89303 | invasive | non-epithelial | endometrial stromal sarcoma, nos |
| 89313 | invasive | non-epithelial | endometrial stromal sarcoma, low grade |
| 89333 | invasive | non-epithelial | adenosarcoma |
| 89363 | invasive | non-epithelial | gastrointestinal stromal sarcoma |
| 89403 | invasive | unknown | mixed tumour malignant |
| 89503 | invasive | epithelial (other) | mullerian mixed tumor |
| 89513 | invasive | epithelial (other) | mesodermal mixed tumor |
| 89803 | invasive | epithelial (other) | carcinosarcoma, nos |
| 89903 | invasive | non-epithelial | mesenchymoma malignant |
| 90143 | invasive | epithelial (serous) | serous adenocarcinofibroma |
| 90603 | invasive | non-epithelial (germ cell) | dysgerminoma |
| 90643 | invasive | non-epithelial (germ cell) | germinoma |
| 90713 | invasive | non-epithelial (germ cell) | endodermal sinus tumour |
| 90803 | invasive | non-epithelial (germ cell) | teratoma, malignant nos |
| 90813 | invasive | non-epithelial (germ cell) | teratocarcinoma |
| 90833 | invasive | non-epithelial (germ cell) | malignant teratoma, intermediate |
| 90843 | invasive | non-epithelial (germ cell) | dermoid cyst with mal transform |
| 90853 | invasive | non-epithelial (germ cell) | mixed germ cell tumor |
| 90903 | invasive | non-epithelial (germ cell) | struma ovarii, malignant |
| 91103 | invasive | epithelial (other) | mesonephroma malignant |
| 93643 | invasive | other | ewing sarcoma / pnet |
| 95003 | invasive | other | neuroblastoma |
