## Supplementary material for "CA125 and age-based models for ovarian cancer detection in primary care: a population-based external validation study": Ovatools risk prediction formulae

```
****SUPPLEMENT 2
****This code was applied using Stata version 18.

/*
CALCULATE THE RISK OF INVASIVE OVARIAN & OVARIAN FOR EACH PARTICIPANT BASED ON
THE  PREDICTIVE MODEL FORMULA USING CA125 AND AGE

ORIGINAL KNOTS:
Log ca125                       -1.390562 -0.8027754 -0.5150933 -0.1667867
0.7376696
Age mean centred                -24 -9 -1 10 27

*/
cd "G:\Primary Care\OVATOOLS\Data\Data prep"
use cohort, clear
keep epatid CA125 test age ovaryca invasive age_cat

***create new splines for new dataset:
*log transform CA125, floor & mc
gen log_ca125=log(CA125)
sum log_ca125, d // mean 2.569051
*replace log_ca125=log_ca125-2.569051
replace log_ca125=log_ca125-3
sum log_ca125, d

*mean centre and floor age
sum age, d // mean 54.71233
replace age=floor(age)
gen agemc=age-55
order epatid CA125 age agemc
sum agemc, d
sort age CA125

*gen splines for new dataset
mkspline log_ca125_=log_ca125, cubic knots (-1.390562 -0.8027754 -0.5150933
-0.1667867 0.7376696)
matrix K=r(knots)

mkspline age_=agemc, cubic knots (-24 -9 -1 10 27)
matrix KA=r(knots)

********************************************************************************
********************************************************************************
*CALCUATE INDIVIDUAL RISK FOR EACH MODEL
*OVARIAN CANCER MODEL
gen log_odds_ovary=ln(0.0002362) +(log_ca125_1*ln(2.26625))
+(log_ca125_2*ln(0.0046567)) +(log_ca125_3*ln(1.25*10^31))
+(log_ca125_4*ln(3.40*10^-56)) + (age_1*ln(0.9263145)) + (age_2*ln(1.746288)) +
(age_3*ln(0.1047997)) + (age_4*ln(7.06957))

*INVASIVE-OVARIAN CANCER MODEL
gen log_odds_invasive=ln(0.0000571) +(log_ca125_1*ln(1.069068))
+(log_ca125_2*ln(0.0007832)) +(log_ca125_3*ln(8.33*10^43))
+(log_ca125_4*ln(3.36*10^-79)) + (age_1*ln(0.9415075)) + (age_2*ln(1.829243)) +
```

```
(age_3*ln(0.0776767)) + (age_4*ln(10.23064))

*convert log odds to probability [0,1]
gen risk_ovary=exp(log_odds_ovary)/(1+exp(log_odds_ovary))

*invasive ovarian cancer risk
gen risk_invasive=exp(log_odds_invasive)/(1+exp(log_odds_invasive))

save risk_all_models, replace
```
