## Supplementary material for "CA125 and age-based models for ovarian cancer detection in primary care: a population-based external validation study": CA125 levels equating 1% and 3% risk by age group

### Supplement 3: Average CA125 levels equating to approximately 1-2.9% and ≥3% risk of invasive ovarian cancer by age group using the Ovatools model

| Age group | Equivalent CA125 level (U/ml) | |
|---|---|---|
| | Ovatools 1-2.9% risk | Ovatools ≥3% risk |
| 18-29 years | 34-90.9 | ≥91 |
| 30-39 years | 59-159.9 | ≥160 |
| 40-49 years | 58-156.9 | ≥157 |
| 50-59 years | 26-56.9 | ≥57 |
| 60-69 years | 22-36.9 | ≥37 |
| 70-79 years | 22-40.9 | ≥41 |
| 80-89 years | 26-57.9 | ≥58 |
