## Additional performance metrics - by risk level, demographics and any OC for "CA125 and age-based models for ovarian cancer detection in primary care: a population-based external validation study"

### Supplement 4: Additional model validation metrics

#### 4.1. Performance by risk level

Participants were ranked by ascending predicted risk level and grouped into bins of 5000. The mean predicted risk was plotted against the mean outcome for each bin (each bin is represented by a small blue dot in the graph). Predicted risk levels above 5% are not displayed because they were considered outliers, are not clinically relevant and formed a very small proportion of the population sample (1.18%). For lower risk predictions, the model slightly under predicted risk, and for higher predictions the model overpredicted risk. Agreement between predicted and observed outcomes was greater for lower prediction levels.

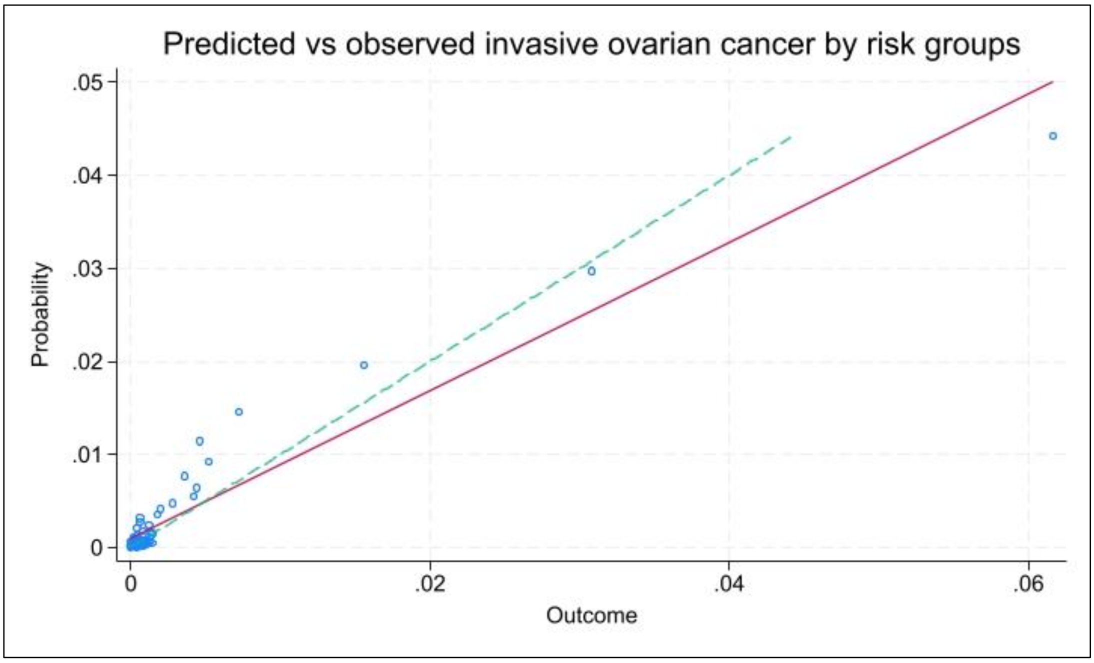

##### 4.2. Invasive ovarian cancer model performance by ethnicity, deprivation and early-stage invasive ovarian cancer

| | | AUC | Intercept | Slope | O:E | CITL |
|---|---|---|---|---|---|---|
| Early stage invasive OC (excludes missing & stage III-IV) | | 0.885 (0.867-0.902) | 0.0038 (0.0037-0.0040) | 0.751 (0.751-0.751) | 0.333 | -1.344 |
| Ethnicity group | Asian/Asian British | 0.899 (0.849-0.950) | 0.0007(0.0000-0.0015) | 0.900 (0.899-0.901) | 0.896 | -0.140 |
| | Black/Black British | 0.953 (0.900-1.000) | 0.0015 (0.0009-0.0022) | 1.119 (1.118-1.120) | 0.554 | -0.745 |
| | Mixed | 0.944 (0.894-0.993) | 0.0017 (0.0011-0.0024) | 0.989 (0.988-0.990) | 0.603 | -0.675 |
| | White/White British | 0.947 (0.940-0.954) | 0.0003 (0.0001-0.0005) | 1.004 (1.004-1.004) | 0.958 | -0.063 |
| | Other | 0.936 (0.857-1.000) | 0.0003 (-0.0008-0.0014) | 1.114 (1.113-1.115) | 0.827 | -0.239 |
| Deprivation quintiles (1=least, 5=most deprived) | Quintile 1 | 0.947(0.934-0.960) | 0.0003 ((-0.0001)-0.0007) | 1.008 (1.008-1.008) | 0.952 | -0.073 |
| | Quintile 2 | 0.953 (0.941-0.965) | 0.0003 ((-0.0001)-0.0008) | 1.005 (1.005-1.005) | 0.948 | -0.078 |
| | Quintile 3 | 0.940 (0.926-0.955) | 0.0004 ((-0.0001)-0.0008) | 0.977 (0.977-0.977) | 0.972 | -0.041 |
| | Quintile 4 | 0.942 (0.925-0.958) | 0.0003 ((-0.0002)-0.0007) | 1.005 (1.005-1.005) | 0.952 | -0.069 |
| | Quintile 5 | 0.945 (0.926-0.965) | 0.0011 (0.0007-0.0015) | 1.030 (1.030-1.030) | 0.775 | -0.334 |
| AUC=area under the curve; CITL = calibration in the large; OC = ovarian cancer; O:E=observed vs expected outcomes | | | | | | |

##### 4.3. Any ovarian cancer model performance for all participants

| AUC (95% CI) | Intercept (95% CI) | Slope (95% CI) | O:E | CITL |
|---|---|---|---|---|
| 0.925 (0.918-0.931) | 0.0009 (0.0007-0.0011) | 1.010 (1.010-1.010) | 0.889 | -0.159 |
| AUC = area under the curve, CITL = calibration-in-the-large, O:E = observed versus expected | | | | |

##### 4.4. Invasive ovarian cancer model performance for all participants using age group and CA125 level

| All group | Performance measurement (95% confidence interval) | | | | |
|---|---|---|---|---|---|
| | AUC | Calibration slope | Calibration intercept | CITL | O:E |
| All ages, 18-89 years | 0.947 (0.940-0.954) | 1.001 (0.994-1.008) | 0.0015 ((-0.0054)-0.0083) | -0.111 | 0.926 |
| All ≥50 years | 0.951 (0.943-0.958) | 0.992 (0.983-1.001) | 0.0098 (0.0005-0.0192) | -0.119 | 0.925 |
| All <50 years | 0.897 (0.873-0.922) | 1.073 (1.070-1.076) | 0.0022 ((-0.0009)-0.0052) | -0.077 | 0.932 |
| 18-29 years | 0.804 (0.689-0.920) | 0.789 (0.787-0.791) | 0.0015 ((-0.0009)-0.0038) | -0.683 | 0.520 |
| 30-39 years | 0.885 (0.830-0.941) | 0.942 (0.940- 0.944) | 0.0014 ((-0.0011)-0.0039) | -0.319 | 0.738 |
| 40-49 years | 0.912 (0.885-0.939) | 1.144 (1.141-1.147) | 0.0026 ((-0.0008)-0.0061) | 0.118 | 1.110 |
| 50-59 years | 0.922 (0.900-0.943) | 1.042 (1.036-1.048) | 0.0056 ((-0.0005)-0.0116) | -0.076 | 0.945 |
| 60-69 years | 0.967 (0.957-0.976) | 1.000 (0.990-1.010) | 0.0112 (0.0013-0.0211) | -0.140 | 0.918 |
| 70-79 years | 0.959 (0.948-0.969) | 0.970 (0.957-0.983) | 0.0141 (0.0015-0.0267) | -0.112 | 0.932 |
| 80-89 years | 0.936 (0.915-0.957) | 0.931 (0.917-0.945) | 0.0138 (0.0001-0.0276) | -0.158 | 0.899 |
| AUC=area under the curve; CITL = calibration in the large; O:E=observed vs expected outcomes | | | | | |

##### 4.5. The diagnostic accuracy of using CA125 thresholds equating to ~1% and ~3% Ovatools risk by age group compared to using CA125 ≥35U/ml to detect invasive ovarian cancer

| Age group (invasive ovarian cancer incidence, %) | CA125 threshold (U/ml) | Sensitivity, % (95% CI) | Specificity, % (95% CI) | PPV, % (95% CI) | NPV, % (95% CI) |
|---|---|---|---|---|---|
| 18- 29 years (0.13%) | ≥35 | 56.5 (34.5; 76.8) | 94.8 (94.4; 95.1) | 1.3 (0.7; 2.3) | 99.9 (99.9; 100) |
| | ≥34 | 56.5 (34.5; 76.8) | 94.4 (94.1; 94.8) | 1.3 (0.7; 2.1) | 99.9 (99.9; 100) |
| | ≥91 | 30.4 (13.2; 52.9) | 98.8 (98.6; 98.9) | 3.0 (1.2; 6.1) | 99.9 (99.9; 99.9) |
| 30-39 years (0.14%) | ≥35 | 69.1 (55.2; 80.9) | 92.4 (92.1; 92.6) | 1.2 (0.9; 1.7) | 100 (99.9; 100) |
| | ≥59 | 50.9 (37.1; 64.6) | 97.3 (97.1; 97.5) | 2.6 (1.7; 3.7) | 99.9 (99.9; 100) |
| | ≥160 | 29.1 (17.6; 42.9) | 99.4 (99.3; 99.5) | 6.2 (3.6; 9.9) | 99.9 (99.9; 99.9) |
| 40-49 years (0.26%) | ≥35 | 78.7 (72.7; 83.9) | 92.0 (91.9; 92.2) | 2.5 (2.1; 2.9) | 99.9 (99.9; 100) |
| | ≥58 | 67.4 (60.8; 73.6) | 97.2 (97.1; 97.3) | 5.8 (4.9; 6.8) | 99.9 (99.9; 99.9) |
| | ≥157 | 48.4 (41.7; 55.2) | 99.4 (99.4; 99.5) | 17.4 (14.5; 20.7) | 99.9 (99.9; 99.9) |
| Age 50-59 years (0.52%) | ≥35 | 80.5 (76.3; 84.3) | 95.7 (76.3; 84.3) | 8.8 (7.9; 9.7) | 99.9 (99.9; 99.9) |
| | ≥26 | 84.8 (80.9; 88.2) | 91.6 (91.4; 91.8) | 5.0 (4.5; 5.5) | 99.9 (99.9; 99.9) |
| | ≥57 | 72.3 (67.7; 76.6) | 98.1 (98.0; 98.2) | 16.7 (14.9; 18.5) | 99.9 (99.8; 99.9) |
| Age 60-69 years (1.05%) | ≥35 | 86.9 (83.9; 89.5) | 95.9 (95.8; 96.1) | 18.5 (17.1; 19.9) | 99.9 (99.8; 99.9) |
| | ≥22 | 92.4 (90.0; 94.4) | 89.3 (89.0; 89.5) | 8.4 (7.7; 9.1) | 99.9 (99.8; 99.9) |
| | ≥37 | 86.6 (83.6; 89.2) | 96.2 (96.1; 96.4) | 19.7 (18.2; 21.3) | 99.9 (99.8; 99.9) |
| Age 70-79 years (1.32%) | ≥35 | 87.7 (84.6; 90.3) | 93.6 (93.4; 93.8) | 15.5 (14.2; 16.8) | 99.8 (99.8; 99.9) |
| | ≥22 | 93.5 (91.0; 95.4) | 84.7 (84.4; 85.1) | 7.6 (6.9; 8.2) | 99.9 (99.9; 99.9) |
| | ≥41 | 86.4 (83.2; 89.2) | 94.9 (94.6; 95.1) | 18.3 (16.8; 19.9) | 99.8 (99.8; 99.8) |
| Age 80-89 years (1.26%) | ≥35 | 91.4 (87.7; 94.3) | 87.3 (86.9; 87.8) | 8.7 (7.7; 9.7) | 99.9 (99.8; 99.9) |
| | ≥26 | 92.2 (88.1; 95.1) | 81.8 (81.2; 82.3) | 6.1 (5.3; 6.9) | 99.9 (99.8; 99.9) |
| | ≥58 | 83.1 (78.0; 87.5) | 94.0 (93.6; 94.3) | 15.0 (13.2; 16.9) | 99.8 (99.7; 99.8) |

CA125 = cancer antigen 125, NPV = negative predictive value, PPV = positive predictive value
