## Supplementary material for "CA125 and age-based models for ovarian cancer detection in primary care: a population-based external validation study": Calculating the clinical utility of Ovatools

### Supplement 5: Clinical utility

We report below on the estimated number of women under 90 years who would be referred for further investigation following CA125 in England per year and the number of cases of invasive ovarian cancer (OC) that could be identified using:

(1) Using the Ovatools risk prediction model, where 1-2.9% risk and ≥3% risk triggering primary care ultrasound and direct urgent cancer pathway referral, respectively (**Table S5.1.11.**)

(2) Using integer CA125 thresholds that equate to approximately ≥1% and ≥3% risk of invasive OC by age group to trigger ultrasound and direct cancer referral, respectively. We use age categorisations: 18-29, 30-39, 40-49, 50-59, 60-69, 70-79 and 80-89 years (**Table S5.2.12**)

Utility for both groups is compared to that using current NICE guidelines, in which a CA125 test result ≥35U/ml triggers primary care ultrasound (referred to as "current practice"). For both groups above, we estimate the utility overall, by ages above and below 50 years, and by age group.

**Table S5.i. The number of women tested using CA125 in primary care in England in 2022**

| | Number |
|---|---|
| Women tested with CA125 in 2022 | 34,075 |
| GP practices with CA125 records in CPRD | 1,009 |
| Average women tested using CA125 per GP practice per year | 33.77 |
| Total GP practices in England in 2022[25] | 6,422 |
| Total tested with CA125 in England in 2022 under 90 years | 216,878 |
| Number with invasive ovarian cancer (incidence 0.62%) in England | 1,339 |
| Total population in England in 2022[26] | 57,112,500 |
| CA125 tests per 100,000 population per year | 380 |
| Number with invasive ovarian cancer / 100,000 population | 2.39 |

The mean number of women who received one or more CA125 tests per GP practice per year in CPRD between 28 February 2021 and 1 March 2022 was calculated (**Table S5.i**). Women with a prior diagnosis of any invasive ovarian cancer were excluded. Based on 34,075 women under 90 years tested with CA125 in one year across 1,009 GP practices in CPRD, this equated to approximately 33.77 women per GP practice per year. With 6,422 GP practices in 2022[25], a total of 216,878 women under 90 years were estimated to have been tested with CA125 in England in 2022. The population in England in 2022 was approximately 57,112,500[26], thus, an estimated 380 women had one or more CA125 tests per 100,000 population in 2022, of which 2.39 had invasive ovarian cancer.

**Table S5.ii. The number of women under 90 years tested using CA125 in primary care in England in 2022 by age group**

| Age group | Population distribution, % | Number tested using CA125 per year | Invasive OC incidence, n (%) |
|---|---|---|---|
| 18-29 years | 5.40% | 11,711 | 23 (0.13) |
| 30-39 years | 11.69% | 25,353 | 55 (0.14) |
| 40-49 years | 25.17% | 54,588 | 221 (0.26) |
| 50-59 years | 22.91% | 49,687 | 401 (0.52) |
| 60-69 years | 16.88% | 36,609 | 603 (1.05) |
| 70-79 years | 11.98% | 25,982 | 536 (1.32) |
| 80-89 years | 5.97% | 12,948 | 255 (1.26) |

### Section 5.1. Clinical utility of using Ovatools risk models at 1-2.9% and ≥3% to trigger ultrasound and urgent referral, respectively, compared to current practice.

**Table S5.1.1. Distribution of CA125 ≥35U/ml and Ovatools risk among CA125-tested women per year, all ages**

| CA125 and Ovatools risk distribution for all participants and by invasive ovarian cancer diagnosis | | Proportion, % | Number in cohort, total=339,124 | Number in England per year, total=216,878 |
|---|---|---|---|---|
| All participants | CA125 <35U/mL | 93.20 | 316,068 | 202,133 |
| | CA125 ≥35U/mL | 6.80 | 23,056 | 14,745 |
| | <1% risk | 92.05 | 312,152 | 199,629 |
| | ≥1% risk | 7.95 | 26,972 | 17,249 |
| | 1-3% | 5.25 | 17,806 | 11,387 |
| | ≥3% risk | 2.70 | 9,166 | 5,862 |
| Participants with invasive ovarian cancer | CA125 ≥35U/mL | 84.67 | 1,773 | 1,134 |
| | ≥1% risk | 86.82 | 1,818 | 1,163 |
| | ≥3% risk | 77.55 | 1,624 | 1,039 |
| Participants without invasive ovarian cancer | CA125 ≥35U/mL | 6.31 | 21,283 | 13,611 |
| | ≥1% risk | 7.46 | 25,154 | 16,087 |
| | ≥3% risk | 2.24 | 7,542 | 4,238 |

**Table S5.1.2. Distribution of CA125 ≥35U/ml and Ovatools risk among CA125-tested women per year, 18-29 years**

| CA125 and Ovatools risk distribution for all participants and by invasive ovarian cancer diagnosis | | Proportion, % | Number in cohort, total=18,319 | Number in England per year, total=11,711 |
|---|---|---|---|---|
| All participants | CA125 <35U/mL | 94.69 | 17,347 | 11,090 |
| | CA125 ≥35U/mL | 5.31 | 972 | 621 |
| | <1% risk | 95.39 | 17,474 | 11,171 |
| | ≥1% risk | 4.61 | 845 | 540 |
| | 1-3% | 3.53 | 647 | 414 |
| | ≥3% risk | 1.08 | 198 | 127 |
| Participants with invasive ovarian cancer | CA125 ≥35U/mL | 56.52 | 13 | 8 |
| | ≥1% risk | 47.83 | 11 | 7 |
| | ≥3% risk | 39.13 | 9 | 6 |
| Participants without invasive ovarian cancer | CA125 ≥35U/mL | 5.24 | 959 | 613 |
| | ≥1% risk | 4.56 | 834 | 533 |
| | ≥3% risk | 1.03 | 189 | 118 |

**Table S5.1.3. Distribution of CA125 ≥35U/ml and Ovatools risk among CA125-tested women per year, 30-39 years**

| CA125 and Ovatools risk distribution for all participants and by invasive ovarian cancer diagnosis | | Proportion, % | Number in cohort, total=39,635 | Number in England per year, total=25,353 |
|---|---|---|---|---|
| All participants | CA125 <35U/mL | 0.14 | 55 | 35 |
| | CA125 ≥35U/mL | 99.86 | 39,580 | 25,318 |
| | <1% risk | 92.27 | 36,570 | 23,392 |
| | ≥1% risk | 7.73 | 3,065 | 1,961 |
| | 1-3% | 97.43 | 38,617 | 24,702 |
| | ≥3% risk | 2.57 | 1,018 | 651 |
| Participants with invasive ovarian cancer | CA125 ≥35U/mL | 69.09 | 38 | 24 |
| | ≥1% risk | 49.09 | 27 | 17 |
| | ≥3% risk | 23.64 | 13 | 8 |
| Participants without invasive ovarian cancer | CA125 ≥35U/mL | 7.65 | 3,027 | 1,936 |
| | ≥1% risk | 2.50 | 991 | 634 |
| | ≥3% risk | 0.55 | 219 | 135 |

**Table S5.1.4. Distribution of CA125 ≥35U/ml and Ovatools risk among CA125-tested women per year, 40-49 years**

| CA125 and Ovatools risk distribution for all participants and by invasive ovarian cancer diagnosis | | Proportion, % | Number in cohort, total=39,635 | Number in England per year, total=25,353 |
|---|---|---|---|---|
| All participants | CA125 <35U/mL | 91.86 | 78,396 | 50,144 |
| | CA125 ≥35U/mL | 8.14 | 6,948 | 4,444 |
| | <1% risk | 96.82 | 82,628 | 52,851 |
| | ≥1% risk | 3.18 | 2,716 | 1,737 |
| | 1-3% | 2.44 | 2,084 | 1,333 |
| | ≥3% risk | 0.74 | 632 | 404 |
| Participants with invasive ovarian cancer | CA125 ≥35U/mL | 78.73 | 174 | 111 |
| | ≥1% risk | 66.52 | 147 | 94 |
| | ≥3% risk | 51.13 | 113 | 72 |
| Participants without invasive ovarian cancer | CA125 ≥35U/mL | 7.96 | 6,774 | 4,333 |
| | ≥1% risk | 3.02 | 2,569 | 1,643 |
| | ≥3% risk | 0.61 | 519 | 291 |

**Table S5.1.5. Distribution of CA125 ≥35U/ml and Ovatools risk among CA125-tested women per year, 50-59 years**

| CA125 and Ovatools risk distribution for all participants and by invasive ovarian cancer diagnosis | | Proportion, % | Number in cohort, total= 77,697 | Number in England per year, total=49,687 |
|---|---|---|---|---|
| All participants | CA125 <35U/mL | 0.52 | 401 | 256 |
| | CA125 ≥35U/mL | 99.48 | 77,296 | 49,430 |
| | <1% risk | 95.26 | 74,012 | 47,330 |
| | ≥1% risk | 4.74 | 3,685 | 2,357 |
| | 1-3% | 0.52 | 401 | 256 |
| | ≥3% risk | 99.48 | 77,296 | 49,430 |
| Participants with invasive ovarian cancer | CA125 ≥35U/mL | 80.55 | 323 | 207 |
| | ≥1% risk | 84.54 | 339 | 217 |
| | ≥3% risk | 72.32 | 290 | 185 |
| Participants without invasive ovarian cancer | CA125 ≥35U/mL | 4.35 | 3,362 | 2,150 |
| | ≥1% risk | 7.03 | 5,434 | 3,475 |
| | ≥3% risk | 1.58 | 1,225 | 679 |

**Table S5.1.6. Distribution of CA125 ≥35U/ml and Ovatools risk among CA125-tested women per year, 60-69 years**

| CA125 and Ovatools risk distribution for all participants and by invasive ovarian cancer diagnosis | | Proportion, % | Number in cohort, total= 57,257 | Number in England per year, total=36,609 |
|---|---|---|---|---|
| All participants | CA125 <35U/mL | 95.05 | 54,420 | 34,795 |
| | CA125 ≥35U/mL | 4.95 | 2,837 | 1,814 |
| | <1% risk | 88.44 | 50,637 | 32,376 |
| | ≥1% risk | 11.56 | 6,620 | 4,233 |
| | 1-3% | 6.93 | 3,969 | 2,538 |
| | ≥3% risk | 4.63 | 2,651 | 1,695 |
| Participants with invasive ovarian cancer | CA125 ≥35U/mL | 86.90 | 524 | 335 |
| | ≥1% risk | 92.37 | 557 | 356 |
| | ≥3% risk | 86.57 | 522 | 334 |
| Participants without invasive ovarian cancer | CA125 ≥35U/mL | 4.08 | 2,313 | 1,479 |
| | ≥1% risk | 10.70 | 6,063 | 3,877 |
| | ≥3% risk | 3.76 | 2,129 | 1,173 |

**Table S5.1.7. Distribution of CA125 ≥35U/ml and Ovatools risk among CA125-tested women per year, 70-79 years**

| CA125 and Ovatools risk distribution for all participants and by invasive ovarian cancer diagnosis | | Proportion, % | Number in cohort, total= 40,624 | Number in England per year, total=25,982 |
|---|---|---|---|---|
| All participants | CA125 <35U/mL | 92.53 | 37,589 | 24,041 |
| | CA125 ≥35U/mL | 7.47 | 3,035 | 1,941 |
| | <1% risk | 84.86 | 34,472 | 22,047 |
| | ≥1% risk | 15.14 | 6,152 | 3,935 |
| | 1-3% | 9.00 | 3,656 | 2,338 |
| | ≥3% risk | 6.14 | 2,496 | 1,596 |
| Participants with invasive ovarian cancer | CA125 ≥35U/mL | 87.69 | 470 | 301 |
| | ≥1% risk | 93.66 | 502 | 321 |
| | ≥3% risk | 86.01 | 461 | 295 |
| Participants without invasive ovarian cancer | CA125 ≥35U/mL | 6.40 | 2,565 | 1,641 |
| | ≥1% risk | 14.09 | 5,650 | 3,614 |
| | ≥3% risk | 5.08 | 2,035 | 1,135 |

**Table S5.1.8. Distribution of CA125 ≥35U/ml and Ovatools risk among CA125-tested women per year, 80-89 years**

| CA125 and Ovatools risk distribution for all participants and by invasive ovarian cancer diagnosis | | Proportion, % | Number in cohort, total= 20,248 | Number in England per year, total=12,948 |
|---|---|---|---|---|
| All participants | CA125 <35U/mL | 87.58 | 17,734 | 11,340 |
| | CA125 ≥35U/mL | 12.42 | 2,514 | 1,608 |
| | <1% risk | 81.00 | 16,400 | 10,487 |
| | ≥1% risk | 19.00 | 3,848 | 2,461 |
| | 1-3% | 11.88 | 2,406 | 1,539 |
| | ≥3% risk | 7.12 | 1,442 | 922 |
| Participants with invasive ovarian cancer | CA125 ≥35U/mL | 90.59 | 231 | 148 |
| | ≥1% risk | 92.16 | 235 | 150 |
| | ≥3% risk | 84.71 | 216 | 138 |
| Participants without invasive ovarian cancer | CA125 ≥35U/mL | 11.42 | 2,283 | 1,460 |
| | ≥1% risk | 18.07 | 3,613 | 2,310 |
| | ≥3% risk | 6.13 | 1,226 | 706 |

**Table S5.1.9. Distribution of CA125 ≥35U/ml and Ovatools risk among CA125-tested women per year, 18-49 years**

| CA125 and Ovatools risk distribution for all participants and by invasive ovarian cancer diagnosis | | Proportion, % | Number in cohort, total= 143,298 | Number in England per year, total=91,653 |
|---|---|---|---|---|
| All participants | CA125 <35U/mL | 92.33 | 132,313 | 84,627 |
| | CA125 ≥35U/mL | 7.67 | 10,985 | 7,026 |
| | <1% risk | 96.80 | 138,719 | 88,724 |
| | ≥1% risk | 3.20 | 4,579 | 2,929 |
| | 1-3% | 2.45 | 3,517 | 2,249 |
| | ≥3% risk | 0.74 | 1,062 | 679 |
| Participants with invasive ovarian cancer | CA125 ≥35U/mL | 75.25 | 225 | 144 |
| | ≥1% risk | 61.87 | 185 | 118 |
| | ≥3% risk | 45.15 | 135 | 86 |
| Participants without invasive ovarian cancer | CA125 ≥35U/mL | 7.52 | 10,760 | 6,882 |
| | ≥1% risk | 3.07 | 4,394 | 2,810 |
| | ≥3% risk | 0.65 | 927 | 544 |

**Table S5.1.10. Distribution of CA125 ≥35U/ml and Ovatools risk among CA125-tested women per year, 50-89 years**

| CA125 and Ovatools risk distribution for all participants and by invasive ovarian cancer diagnosis | | Proportion, % | Number in cohort, total= 143,298 | Number in England per year, total=91,653 |
|---|---|---|---|---|
| All participants | CA125 <35U/mL | 93.84 | 183,755 | 117,506 |
| | CA125 ≥35U/mL | 6.16 | 12,071 | 7,719 |
| | <1% risk | 88.87 | 174,038 | 111,292 |
| | ≥1% risk | 11.44 | 22,393 | 14,320 |
| | 1-3% | 7.30 | 14,289 | 9,137 |
| | ≥3% risk | 4.14 | 8,104 | 5,182 |
| Participants with invasive ovarian cancer | CA125 ≥35U/mL | 86.24 | 1,548 | 990 |
| | ≥1% risk | 90.97 | 1,633 | 1,044 |
| | ≥3% risk | 82.95 | 1,489 | 952 |
| Participants without invasive ovarian cancer | CA125 ≥35U/mL | 5.42 | 10,523 | 6,729 |
| | ≥1% risk | 10.70 | 20,760 | 13,275 |
| | ≥3% risk | 3.41 | 6,615 | 3,693 |

**Table S5.1.11. Women investigated further and identified with invasive OC using current practice compared to Ovatools risks 1-2.9% and ≥3% to trigger ultrasound or cancer referral, respectively.**

| Age group | Age distribution, % | Threshold for primary care ultrasound, U/ml | Ultrasound following CA125, n | Change in USS using Ovatools, n | Urgent referral (risk ≥3%), n | Additional women tested further using Ovatools, n | Invasive OC detected, n | Additional OC detected using Ovatools, n |
|---|---|---|---|---|---|---|---|---|
| All ages | NA | CA125 ≥35U/mL | 14,745 | NA | NA | NA | 1,134 | 29 |
| | | Ovatools 1-2.9% | 11,387 | -3,357 | 5,862 | 2,504 | 1,163 | |
| All ages 18-49 years | 42.26 | CA125 ≥35U/mL | 7,026 | NA | NA | NA | 144 | -26 |
| | | Ovatools 1-2.9% | 2,249 | -4,776 | 679 | -4,097 | 118 | |
| All ages 50-89 years | 57.74 | CA125 ≥35U/mL | 7,719 | NA | NA | NA | 990 | 54 |
| | | Ovatools 1-2.9% | 9,137 | 1,418 | 5,182 | 6,601 | 1,044 | |
| 18-29 years | 5.40 | CA125 ≥35U/mL | 621 | NA | NA | NA | 8 | -1 |
| | | Ovatools 1-2.9% | 414 | -208 | 127 | -81 | 7 | |
| 30-39 years | 11.69 | CA125 ≥35U/mL | 1,961 | NA | NA | NA | 24 | -7 |
| | | Ovatools 1-2.9% | 503 | -1,458 | 148 | -1,309 | 17 | |
| 40-49 years | 25.17 | CA125 ≥35U/mL | 4,444 | NA | NA | NA | 111 | -17 |
| | | Ovatools 1-2.9% | 1,333 | -3,111 | 404 | -2,707 | 94 | |
| 50-59 years | 22.91 | CA125 ≥35U/mL | 2,357 | NA | NA | NA | 207 | 10 |
| | | Ovatools 1-2.9% | 2,723 | 366 | 969 | 1,335 | 217 | |
| 60-69 years | 16.88 | CA125 ≥35U/mL | 1,814 | NA | NA | NA | 335 | 21 |
| | | Ovatools 1-2.9% | 2,538 | 724 | 1,695 | 2,419 | 356 | |
| 70-79 years | 11.98 | CA125 ≥35U/mL | 1,941 | NA | NA | NA | 301 | 20 |
| | | Ovatools 1-2.9% | 2,338 | 397 | 1,596 | 1,994 | 321 | |
| 80-89 years | 5.97 | CA125 ≥35U/mL | 1,608 | NA | NA | NA | 148 | 3 |
| | | Ovatools 1-2.9% | 1,539 | -69 | 922 | 853 | 150 | |

### Section 5.2: Clinical utility using age-group based CA125 levels that approximate 1-2.9% and 3% Ovatools risk to trigger ultrasound and urgent referral, respectively.

**Table 5.2.1. The CA125 thresholds used to estimate to trigger ultrasound or cancer referral**

| Age group | Equivalent CA125 level (U/ml) | |
|---|---|---|
| | Ovatools 1-2.9% risk (primary care ultrasound) | Ovatools ≥3% risk (urgent cancer referral) |
| 18-29 years | 34-90.9 | ≥91 |
| 30-39 years | 59-159.9 | ≥160 |
| 40-49 years | 58-156.9 | ≥157 |
| 50-59 years | 26-56.9 | ≥57 |
| 60-69 years | 22-36.9 | ≥37 |
| 70-79 years | 22-40.9 | ≥41 |
| 80-89 years | 26-57.9 | ≥58 |

**Table S5.2.2. Distribution of women with CA125 values above/below the 1% and 3% risk thresholds, all ages**

| CA125 / risk distribution for all participants and by invasive ovarian cancer diagnosis | | Proportion, % | Number in cohort, total=339,124 | Number in England per year, total=216,878 |
|---|---|---|---|---|
| All participants | <1% risk* | 91.08 | 308,877 | 197,534 |
| | ≥1% risk* | 8.92 | 30,247 | 19,344 |
| | 1-3%* | 6.06 | 20,550 | 13,142 |
| | ≥3% risk* | 2.86 | 9,697 | 6,201 |
| Participants with invasive ovarian cancer | ≥1% risk* | 86.82 | 1,818 | 1,163 |
| | ≥3% risk* | 77.55 | 1,624 | 1,039 |
| Participants without invasive ovarian cancer | ≥1% risk* | 7.46 | 25,154 | 16,087 |
| | ≥3% risk* | 2.24 | 7,542 | 4,238 |
| *CA125 thresholds applied by age group as defined in Table S5.2.1 | | | | |

**Table S5.2.3. Distribution of women with CA125 values above/below the 1% and 3% risk thresholds, 18-29 years**

| CA125 /risk distribution for all participants and by invasive ovarian cancer diagnosis, U/ml (% risk) | | Proportion, % | Number in cohort, total=18,319 | Number in England per year, total=11,711 |
|---|---|---|---|---|
| All participants | CA125 <34 (<1%) | 94.37 | 17,287 | 11,052 |
| | CA125 = 34-90.9 (1-3% risk) | 4.36 | 799 | 511 |
| | CA125 ≥91 (≥3% risk) | 1.27 | 233 | 149 |
| Participants with invasive ovarian cancer | CA125 ≥34 (≥1% risk) | 56.52 | 13 | 8 |
| | CA125 ≥91 (≥3% risk) | 30.43 | 7 | 4 |
| Participants without invasive ovarian cancer | CA125 ≥34 (≥1% risk) | 7.46 | 5.57 | 1,019 |
| | CA125 ≥91 (≥3% risk) | 2.24 | 1.24 | 226 |

**Table S5.2.4. Distribution of women with CA125 values above/below the 1% and 3% risk thresholds, 30-39 years**

| CA125 /risk distribution for all participants and by invasive ovarian cancer diagnosis, U/ml (% risk) | | Proportion, % | Number in cohort, total=39,635 | Number in England per year, total=25,353 |
|---|---|---|---|---|
| All participants | CA125 <59 (<1%) | 97.24 | 38,541 | 24,653 |
| | CA125 = 59-159.9 (1-3% risk) | 2.11 | 837 | 535 |
| | CA125 ≥160 (≥3% risk) | 0.65 | 257 | 164 |
| Participants with invasive ovarian cancer | CA125 ≥59 (≥1% risk) | 50.91 | 28 | 18 |
| | CA125 ≥160 (≥3% risk) | 29.09 | 16 | 10 |
| Participants without invasive ovarian cancer | CA125 ≥59 (≥1% risk) | 2.69 | 1,066 | 682 |
| | CA125 ≥160 (≥3% risk) | 0.61 | 241 | 148 |

**Table S5.2.5. Distribution of women with CA125 values above/below the 1% and 3% risk thresholds, 40-49 years**

| CA125 /risk distribution for all participants and by invasive ovarian cancer diagnosis, U/ml (% risk) | | Proportion, % | Number in cohort, total=85,344 | Number in England per year, total=54,588 |
|---|---|---|---|---|
| All participants | CA125 <58 (<1%) | 97.00 | 82,784 | 52,951 |
| | CA125 = 58-156.9 (1-3% risk) | 2.28 | 1,946 | 1,245 |
| | CA125 ≥157 (≥3% risk) | 0.72 | 614 | 393 |
| Participants with invasive ovarian cancer | CA125 ≥58 (≥1% risk) | 67.42 | 149 | 95 |
| | CA125 ≥157 (≥3% risk) | 48.42 | 107 | 68 |
| Participants without invasive ovarian cancer | CA125 ≥58 (≥1% risk) | 2.83 | 2,411 | 1,542 |
| | CA125 ≥157 (≥3% risk) | 0.60 | 507 | 286 |

**Table S5.2.6. Distribution of women with CA125 values above/below the 1% and 3% risk thresholds, 50-59 years**

| CA125 /risk distribution for all participants and by invasive ovarian cancer diagnosis, U/ml (% risk) | | Proportion, % | Number in cohort, total=77,697 | Number in England per year, total=49,687 |
|---|---|---|---|---|
| All participants | CA125 <26 (<1%) | 91.19 | 70,849 | 45,307 |
| | CA125 = 26-56.9 (1-3% risk) | 6.57 | 75,956 | 3,266 |
| | CA125 ≥57 (≥3% risk) | 2.24 | 1,741 | 1,113 |
| Participants with invasive ovarian cancer | CA125 ≥26 (≥1% risk) | 84.79 | 340 | 217 |
| | CA125 ≥57 (≥3% risk) | 72.32 | 290 | 185 |
| Participants without invasive ovarian cancer | CA125 ≥26 (≥1% risk) | 8.42 | 6,508 | 4,162 |
| | CA125 ≥57 (≥3% risk) | 1.88 | 1,451 | 823 |

**Table S5.2.7. Distribution of women with CA125 values above/below the 1% and 3% risk thresholds, 60-69 years**

| CA125 /risk distribution for all participants and by invasive ovarian cancer diagnosis, U/ml (% risk) | | Proportion, % | Number in cohort, total=57,257 | Number in England per year, total=36,609 |
|---|---|---|---|---|
| All participants | CA125 <22 (<1%) | 88.42 | 50,625 | 32,369 |
| | CA125 = 22-36.9 (1-3% risk) | 6.95 | 54,607 | 2,546 |
| | CA125 ≥37 (≥3% risk) | 4.63 | 2,650 | 1,694 |
| Participants with invasive ovarian cancer | CA125 ≥22 (≥1% risk) | 92.37 | 557 | 356 |
| | CA125 ≥37 (≥3% risk) | 86.57 | 522 | 334 |
| Participants without invasive ovarian cancer | CA125 ≥22 (≥1% risk) | 10.72 | 6,075 | 3,884 |
| | CA125 ≥37 (≥3% risk) | 3.76 | 2,128 | 1,172 |

**Table S5.2.8. Distribution of women with CA125 values above/below the 1% and 3% risk thresholds, 70-79 years**

| CA125 /risk distribution for all participants and by invasive ovarian cancer diagnosis, U/ml (% risk) | | Proportion, % | Number in cohort, total=40,624 | Number in England per year, total=25,982 |
|---|---|---|---|---|
| All participants | CA125 <22 (<1%) | 83.70 | 34,004 | 21,748 |
| | CA125 = 22-40.9 (1-3% risk) | 10.08 | 38,097 | 2,618 |
| | CA125 ≥41 (≥3% risk) | 6.22 | 2,527 | 1,616 |
| Participants with invasive ovarian cancer | CA125 ≥22 (≥1% risk) | 93.47 | 501 | 320 |
| | CA125 ≥41 (≥3% risk) | 86.38 | 463 | 296 |
| Participants without invasive ovarian cancer | CA125 ≥22 (≥1% risk) | 15.26 | 6,119 | 3,914 |
| | CA125 ≥41 (≥3% risk) | 5.15 | 2,064 | 1,153 |

**Table S5.2.9. Distribution of women with CA125 values above/below the 1% and 3% risk thresholds, 80-89 years**

| CA125 /risk distribution for all participants and by invasive ovarian cancer diagnosis, U/ml (% risk) | | Proportion, % | Number in cohort, total=20,248 | Number in England per year, total=12,948 |
|---|---|---|---|---|
| All participants | CA125 <26 (<1%) | 80.84 | 16,369 | 10,467 |
| | CA125 = 26-57.9 (1-3% risk) | 12.17 | 18,833 | 1,576 |
| | CA125 ≥58 (≥3% risk) | 6.99 | 1,415 | 905 |
| Participants with invasive ovarian cancer | CA125 ≥26 (≥1% risk) | 92.16 | 235 | 150 |
| | CA125 ≥58 (≥3% risk) | 83.14 | 212 | 136 |
| Participants without invasive ovarian cancer | CA125 ≥26 (≥1% risk) | 18.23 | 3,644 | 2,330 |
| | CA125 ≥58 (≥3% risk) | 6.02 | 1,203 | 693 |

**Table S5.2.10. Distribution of women with CA125 values above/below the 1% and 3% risk thresholds, 18-49 years**

| CA125 and Ovatools risk distribution for all participants and by invasive ovarian cancer diagnosis | | Proportion, % | Number in cohort, total=143,298 | Number in England per year, total=91,653 |
|---|---|---|---|---|
| All participants | <1% risk* | 96.73 | 138,612 | 88,655 |
| | 1-3%* | 2.50 | 3,582 | 2,291 |
| | ≥3% risk* | 0.77 | 1,104 | 706 |
| Participants with invasive ovarian cancer | ≥1% risk* | 63.55 | 190 | 122 |
| | ≥3% risk* | 43.48 | 130 | 83 |
| Participants without invasive ovarian cancer | ≥1% risk* | 3.14 | 4,496 | 2,876 |
| | ≥3% risk* | 0.68 | 974 | 576 |
| *CA125 thresholds applied by age group as defined in Table S5.2.1 | | | | |

**Table S5.2.11. Distribution of women with CA125 values above/below the 1% and 3% risk thresholds, 50-89 years**

| CA125 and Ovatools risk distribution for all participants and by invasive ovarian cancer diagnosis | | Proportion, % | Number in cohort, total=195,826 | Number in England per year, total=125,225 |
|---|---|---|---|---|
| All participants | <1% risk* | 87.75 | 171,847 | 109,891 |
| | ≥1% risk* | 7.99 | 15,646 | 10,005 |
| | 1-3%* | 4.26 | 8,333 | 5,329 |
| | ≥3% risk* | 2.86 | 9,697 | 6,201 |
| Participants with invasive ovarian cancer | ≥1% risk* | 90.97 | 1,633 | 1,044 |
| | ≥3% risk* | 82.84 | 1,487 | 951 |
| Participants without invasive ovarian cancer | ≥1% risk* | 11.52 | 22,346 | 14,290 |
| | ≥3% risk* | 3.53 | 6,846 | 3,842 |
| *CA125 thresholds applied by age group as defined in Table S5.2.1 | | | | |

**Table S5.2.12. The clinical utility of using CA125 thresholds that approximate 1 and 3% risk by age group to trigger ultrasound and cancer referral, respectively, compared to current practice.**

| Age group | Age distribution, % | Threshold for primary care ultrasound, U/ml (% risk) | Ultrasound following CA125, n | Change in ultrasound using Ovatools, n | Urgent referral (risk ≥3%*), n | Additional tested further using Ovatools, n | Invasive OC detected, n | Additional OC detected using Ovatools, n |
|---|---|---|---|---|---|---|---|---|
| All ages | NA | CA125 ≥35 | 14,745 | NA | NA | NA | 1,134 | 32 |
| | | 1-2.9% risk* | 12,296 | -2449 | 6,035 | 3,586 | 1,166 | |
| All 18-49 years | 42.26 | CA125 ≥35 | 7,026 | NA | NA | NA | 144 | -22 |
| | | 1-2.9% risk* | 2,291 | -4735 | 706 | -4029 | 122 | |
| All 50-89 years | 57.74 | CA125 ≥35 | 7,719 | NA | NA | NA | 990 | 54 |
| | | 1-2.9% risk* | 10,005 | 2286 | 5,329 | 7,615 | 1,044 | |
| 18-29 years | 5.40 | CA125 ≥35 | 621 | NA | NA | NA | 8 | 0 |
| | | CA125 ≥34 (≥1% risk) | 511 | -111 | 149 | 38 | 8 | |
| 30-39 years | 11.69 | CA125 ≥35 | 1,961 | NA | NA | NA | 24 | -6 |
| | | CA125 ≥59 (≥1% risk) | 535 | -1425 | 164 | -1261 | 18 | |
| 40-49 years | 25.17 | CA125 ≥35 | 4,444 | NA | NA | NA | 111 | -16 |
| | | CA125 ≥58 (≥1% risk) | 1,245 | -3199 | 393 | -2807 | 95 | |
| 50-59 years | 22.91 | CA125 ≥35 | 2,357 | NA | NA | NA | 207 | 11 |
| | | CA125 ≥26 (≥1% risk) | 3,266 | 909 | 1113 | 2,023 | 217 | |
| 60-69 years | 16.88 | CA125 ≥35 | 1,814 | NA | NA | NA | 335 | 21 |
| | | CA125 ≥22 (≥1% risk) | 2,546 | 732 | 1,694 | 2,426 | 356 | |
| 70-79 years | 11.98 | CA125 ≥35 | 1,941 | NA | NA | NA | 301 | 20 |
| | | CA125 ≥22 (≥1% risk) | 2,618 | 677 | 1,616 | 2,293 | 320 | |
| 80-89 years | 5.97 | CA125 ≥35 | 1,608 | NA | NA | NA | 148 | 3 |
| | | CA125 ≥26 (≥1% risk) | 1,576 | -32 | 905 | 873 | 150 | |
| *CA125 thresholds equating approximate risk applied by age group as defined in Table S5.2.1 | | | | | | | | |
