## Supplementary material for "CA125 and age-based models for ovarian cancer detection in primary care: a population-based external validation study": Sample size calculation

### Supplement 6: Sample size calculation

Taking the approach of Riley *et al*. (1), we calculated the required sample size for precise estimation of observed divided by expected cases, the calibration slope, the C-statistic and the net benefit (clinical utility) at a referral threshold of $\geq$3%. Riley *et al.* (1) recommend that the sample size is at least as large as the maximum of the four required sample size figures. The inputs to the calculations were derived from the estimation set in the model development study (2) as follows: (i) Proportion with disease = 0.009, (ii) Mean and variance of the linear predictor in those with disease = -2.09 and 2.52, respectively (iii) Mean and variance of the linear predictor in those without disease = -6.34 and 1.32, respectively, (iv) C-statistic = 0.92, however, we used C = 0.82 to allow for shrinkage, (v) Sensitivity and specificity at a threshold of 3% = 0.721 and 0.967, respectively.

We anticipated the observed versus expected (O/E) statistic would close to 1.0, and to obtain 95% confidence intervals (CIs) on the logarithm which would retransform to approximately +/- 0.05, we stipulate a standard error (SE) of the logarithm of O/E of 0.025. For this, we calculated a sample size of 176,178 subjects for external validation. For the calibration slope, the shape of the linear predictor distribution in those with ovarian cancer (OC) appeared approximately normal in the estimation dataset. We anticipated the slope, and intercept would be approximately 1 and zero, respectively. We required a SE on the slope of 0.025, giving a 95% CI of approximately +/- 0.05. The distribution in those without OC appeared approximately lognormal. Applying these to a simulated population of 1,000,000 subjects with a mixing fraction of 0.009 (see above), we required a sample of 140,800 subjects. For the C-statistic of 0.92, disease prevalence of 0.009 and a required SE of 0.025, we needed 13,648 subjects. For the more conservative C-statistic of 0.82, we required 32,012 subjects. From the sensitivities and specificities above, the standardised net benefit in the estimation set was 0.61 to have a 95% CI extending no further than =/- 0.1, we specify a SE of 0.05. To achieve a SE of at most 0.05, we needed 38,228 subjects. Using the maximum of the four figures, 176,178 subjects was the minimum sample size of the validation set. We repeated calculations for the subgroup of invasive OC with prevalence 0.007 and C-statistic in the estimation set of 0.946. We specified the same required SE. The minimum study sizes arrived at were: O/E: 226,968 subjects; Calibration slope: 39,876 subjects; C-statistic: 14,056 for C=0.94, 36,788 for C=0.84; Net benefit: 43,372. Using the maximum of these, we aimed for a sample size of at least 226,968 subjects. In our initial sample size calculations, we used a standard error of 0.05 instead of 0.025 as stipulated above. Thus, we initially calculated that we required 56,742 subjects as the maximum of the four calculations above. However, this mistake in our calculation was rectified, and the final minimum sample size was calculated as 226,968 subjects as demonstrated above.
