## Supplementary material for "CA125 and age-based models for ovarian cancer detection in primary care: a population-based external validation study": Flow diagram demonstrating the application of inclusion and exclusion criteria

### Supplement 8: Applying the inclusion and exclusion criteria

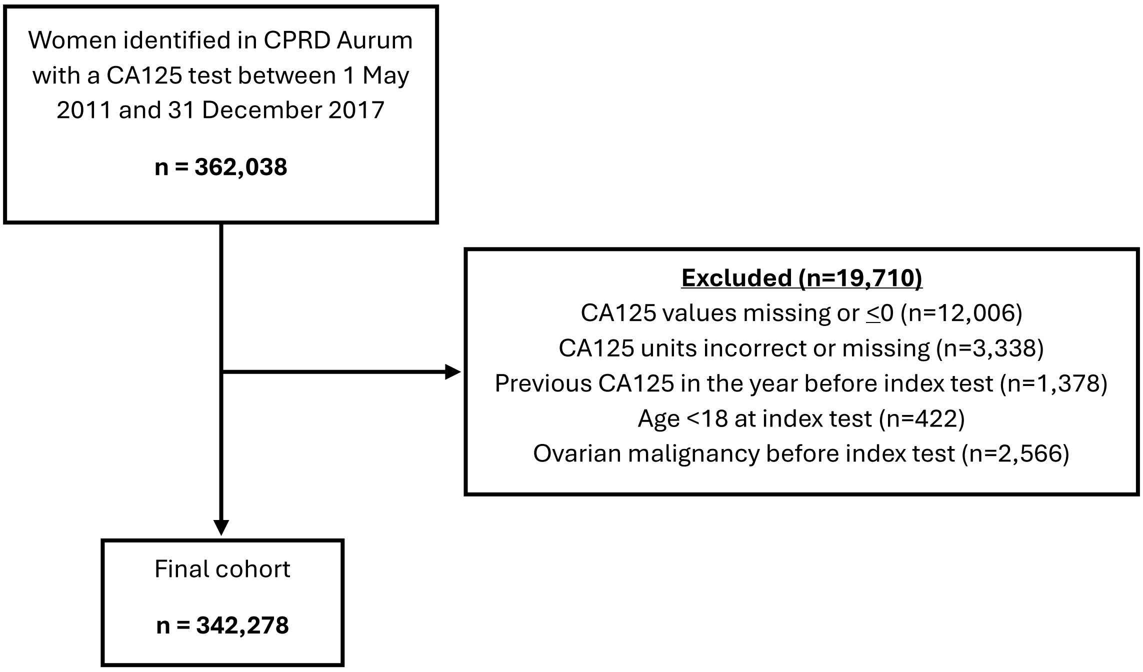


**Flow diagram illustrating the selection of the study cohort and application of exclusion criteria.**
