## Supplementary material for "CA125 and age-based models for ovarian cancer detection in primary care: a population-based external validation study": Ovarian tumour behaviour and morphology

### Supplement 9: Tumour behaviour and morphology

| Histological types | | Participants with any ovarian cancer, n (%) |
|---|---|---|
| Borderline ovarian tumours | | 512 (19.3) |
| Invasive ovarian tumours | | 2143 (80.7) |
| Epithelial | All epithelial | 2,447 (92.2) |
| | Clear cell | 126 (4.8) |
| | Endometrioid | 164 (6.2) |
| | Mucinous | 384 (14.5) |
| | Serous | 1,313 (49.5) |
| | Other | 122 (4.6) |
| | Unknown | 338 (12.7) |
| Non-epithelial | | 99 (3.7) |
| Unknown or other | | 109 (4.1) |
