## Additional diagnostic accuracy metrics - by age group, cancer stage and for any OC for "CA125 and age-based models for ovarian cancer detection in primary care: a population-based external validation study"

### Supplement 10: Additional diagnostic accuracy metrics

**Table S10.1. Diagnostic of accuracy of using CA125 thresholds equating to ~1% and ~3% risk of invasive ovarian cancer, compared to standard practice (CA125 ≥35U/ml)**

| Sub-group (invasive ovarian cancer incidence) | CA125 threshold | Sensitivity, % (95% CI) | Specificity, % (95% CI) | PPV, % (95% CI) | NPV, % (95% CI) |
|---|---|---|---|---|---|
| All ages 18-89 years | ≥35 | 84.9 (83.8; 86.4) | 93.6 (93.5; 93.6) | 7.7 (7.3; 8.0) | 99.9 (99.9; 99.9) |
| | Applied by age category | 87.7 (86.2; 89.1) | 91.6 (91.5; 91.7) | 6.1 (5.8; 6.3) | 99.9 (99.9; 99.9) |
| | | 78.0 (76.1; 79.7) | 97.6 (97.6; 97.6) | 16.8 (16.1; 17.6) | 99.9 (99.8; 99.9) |
| Age 18-49 years (0.21%) | ≥35 | 75.3 (70.0; 80.0) | 92.5 (92.3; 92.6) | 2.0 (1.8; 2.3) | 99.9 (99.9; 100) |
| | ≥46 | 67.9 (62.3; 73.2) | 95.8 (95.7; 95.9) | 3.2 (2.8; 3.7) | 99.9 (99.9; 99.9) |
| | ≥123 | 48.8 (43.0; 54.6) | 99.1 (99.1; 99.2) | 10.7 (9.1; 12.5) | 99.9 (99.9; 99.9) |
| All ages 50-89 years (0.92%) | ≥35 | 86.2 (84.6; 87.8) | 94.6 (94.5; 94.7) | 12.8 (12.2; 13.4) | 99.9 (99.8; 99.9) |
| | Applied by age category | 91.0 (89.6; 92.3) | 88.5 (88.3; 88.6) | 6.8 (6.5; 7.1) | 99.9 (99.9; 99.9) |
| | | 82.8 (81.0; 84.6) | 96.5 (96.4; 96.6) | 17.8 (17.0; 18.7) | 99.8 (99.8; 99.9) |
| Age 50-59 years (0.52%) | ≥35 | 80.5 (76.3; 84.3) | 95.7 (76.3; 84.3) | 8.8 (7.9; 9.7) | 99.9 (99.9; 99.9) |
| | ≥26 | 84.8 (80.9; 88.2) | 91.6 (91.4; 91.8) | 5.0 (4.5; 5.5) | 99.9 (99.9; 99.9) |
| | ≥57 | 72.3 (67.7; 76.6) | 98.1 (98.0; 98.2) | 16.7 (14.9; 18.5) | 99.9 (99.8; 99.9) |
| Age 60-69 years (1.05%) | ≥35 | 86.9 (83.9; 89.5) | 95.9 (95.8; 96.1) | 18.5 (17.1; 19.9) | 99.9 (99.8; 99.9) |
| | ≥22 | 92.4 (90.0; 94.4) | 89.3 (89.0; 89.5) | 8.4 (7.7; 9.1) | 99.9 (99.8; 99.9) |
| | ≥37 | 86.6 (83.6; 89.2) | 96.2 (96.1; 96.4) | 19.7 (18.2; 21.3) | 99.9 (99.8; 99.9) |
| Age 70-79 years (1.32%) | ≥35 | 87.7 (84.6; 90.3) | 93.6 (93.4; 93.8) | 15.5 (14.2; 16.8) | 99.8 (99.8; 99.9) |
| | ≥22 | 93.5 (91.0; 95.4) | 84.7 (84.4; 85.1) | 7.6 (6.9; 8.2) | 99.9 (99.9; 99.9) |
| | ≥41 | 86.4 (83.2; 89.2) | 94.9 (94.6; 95.1) | 18.3 (16.8; 19.9) | 99.8 (99.8; 99.8) |
| Age 80-89 years (1.26%) | ≥35 | 90.6 (88.6; 93.9) | 88.6 (88.1; 89.0) | 9.2 (8.1; 10.4) | 99.9 (99.8; 99.9) |
| | ≥26 | 92.2 (88.1; 95.1) | 81.8 (81.2; 82.3) | 6.1 (5.3; 6.9) | 99.9 (99.8; 99.9) |
| | ≥58 | 83.1 (78.0; 87.5) | 94.0 (93.6; 94.3) | 15.0 (13.2; 16.9) | 99.8 (99.7; 99.8) |

*CA125 = cancer antigen 125, CI = confidence interval; NPV = negative predictive value; PPV = positive predictive value*

**Table S10.2: The diagnostic accuracy of Ovatools at >1% and >3% risk by early and late-stage cancer, compared to using CA125 >35U/ml**

| Outcome (incidence) | CA125/Ovatools threshold | Sensitivity, % (95% CI) | Specificity, % (95% CI) | PPV, % (95% CI) | NPV, % (95% CI) |
|---|---|---|---|---|---|
| Early-stage invasive OC (n=580, 0.17%) | CA125 ≥35U/mL | 66.7 (62.7; 70.6) | 93.6 (93.5; 93.7) | 1.8 (1.6; 1.9) | 99.9 (99.9; 99.9) |
| | ≥1% | 70.7 (66.8; 74.4) | 92.5 (92.4; 92.5) | 1.6 (1.4; 1.7) | 99.9 (99.9; 100) |
| | ≥3% | 51.7 (47.6; 55.9) | 97.8 (97.7; 97.8) | 3.8 (3.4; 4.2) | 99.9 (99.9; 99.9) |
| Late-stage invasive OC (n=1247, 0.37%) | CA125 ≥35U/mL | 93.3 (91.8; 94.7) | 93.6 (93.5; 93.7) | 5.1 (4.8; 5.4) | 100 (100; 100) |
| | ≥1% | 94.6 (93.2; 95.8) | 92.5 (92.4; 92.5) | 4.4 (4.2; 4.7) | 100 (100; 100) |
| | ≥3% | 89.6 (87.7; 91.2) | 97.8 (97.7; 97.8) | 12.8 (12.1; 13.5) | 100 (100; 100) |

**Table S10.3: The diagnostic accuracy of Ovatools at ≥1% and ≥3% risk to detect any ovarian cancer including borderline tumours, compared to using CA125 ≥35U/ml**

| Sub-group (ovarian cancer incidence %) | CA125/Ovatools threshold | Sensitivity, % (95% CI) | Specificity, % (95% CI) | PPV, % (95% CI) | NPV, % (95% CI) |
|---|---|---|---|---|---|
| All women (0.78) | CA125 ≥35U/mL | 78.6 (77.0;80.2) | 93.6 (93.5;93.7) | 8.8 (8.4;9.2) | 99.8 (99.8;99.8) |
| | ≥1% | 81.1 (79.5; 82.5) | 92.5 (92.4;92.5) | 7.7 (7.4; 8.1) | 99.9 (99.8;99.9) |
| | ≥3% | 69.3 (67.5; 71.1) | 97.8 (97.7; 97.8) | 19.5 (18.7; 20.3) | 99.8 (99.7; 99.8) |
| | ≥1% | 88.0 (86.5; 89.3) | 89.2 (89.0; 89.3) | 8.1 (7.8; 8.5) | 99.9 (99.8; 99.9) |
| | ≥3% | 77.8 (76.0; 79.5) | 96.6 (96.5; 96.7) | 19.9 (19.1; 20.8) | 99.7 (99.7; 99.8) |
